# *Persea americana* for Total Health (PATH-2): Effects of Avocado Consumption on Gastrointestinal Health in a Randomized, Crossover, Complete Feeding Trial

**DOI:** 10.64898/2026.03.15.26348343

**Authors:** María G. Sanabria-Véaz, Tori A. Holthaus, Maggie Oleksiak, David Revilla, David A. Alvarado, Marahí Pérez-Tamayo, Naiman A. Khan, Hannah D. Holscher

## Abstract

**Background:** Diets rich in monounsaturated fatty acids (MUFAs) and fiber support gastrointestinal health and the microbiome; however, the effect of whole foods relative to their isolated nutrients remains under-investigated.

**Objective:** Determine the impact of avocado consumption on gastrointestinal health and microbiome beyond the individual effects of MUFAs and fiber.

**Methods:** Adults with overweight and obesity (n=43, mean age=41y, BMI=31.6kg/m^2^) completed a randomized, crossover, controlled feeding study with three 4-wk dietary interventions separated by 2-wk washouts: average American (AA), oleic acid + fiber (OF) nutrients, and avocado (AV). The base diet was supplemented with 209g avocado (AV), or isocaloric snacks high in MUFA/fiber (OF) or low in MUFA/fiber (AA). Outcomes included fecal microbiome (shotgun metagenomics), fecal microbial metabolites (short-chain [SCFA] and branched-chain [BCFA] fatty acids, phenols, indoles, and bile acids), intestinal permeability (24h urinary sweetener excretion), systemic (CRP, IL-6, LBP) and gut (fecal calprotectin and sIgA) inflammatory markers, and gastrointestinal tolerance symptoms. Statistical analysis included linear mixed models, Friedman tests, and multivariable association analysis.

**Results:** Fecal acetate and total SCFAs were 28% and 18% higher in AV and OF conditions, compared to AA (p<0.001 & p=0.019, respectively). Total secondary bile acids in the AV condition were 34% and 24% lower compared to OF (p<0.001) and AA (p=0.011), respectively. *Alistipes communis* (β=0.85, q=0.03) and *Bacteroides uniformis* (β=0.50, q=0.14) were higher following AV, whereas *Lachnospira eligens* (β=1.79, q <0.001) was higher following OF, compared to AA. Microbial genes involved in pectin, cellulose, and hemicellulose degradation were enriched in AV and OF. Fecal calprotectin was lower in AV (30%; p=0.03) and OF (26%; p=0.04) compared to AA, while sIgA was 34% lower following AV, compared to AA (p=0.01).

**Conclusions:** Avocado and MUFA/fiber-matched control had similar fermentation, but distinct secondary bile acid and microbial profiles, emphasizing the food matrix and gut microbiome as key determinants of diet-health relations.

**Clinical Trial Registry number and website where it was obtained:** https://clinicaltrials.gov/study/NCT05941728?intr=NCT05941728&rank=1

## INTRODUCTION

Dietary fiber and fat are key modulators of gastrointestinal and metabolic health, in part through their effects on the gut microbiome (1,2), defined as the collection of species and their genes in the gut (3). Dietary fibers are non-digestible carbohydrates intrinsic and intact in plants or isolated and synthesized that confer health benefits, such as lowering blood glucose and cholesterol, reducing blood pressure, and improving gut transit time and laxation (4,5). Although the adequate intake is ∼28g/day (2), nearly 90% of Americans fail to consume adequate fiber (6). This pattern is consistent with a Western-style diet low in fiber (13-20g/day) (7) and high in saturated fat (8), which has been shown to increase risk for obesity and chronic disease.

The physicochemical properties of fiber influence health benefits and microbial metabolism of fiber in the gut (1,2,7). Soluble and viscous fibers, such as pectin, are readily fermented by intestinal microorganisms, resulting in the production of SCFAs like acetate, butyrate, and propionate, which support energy metabolism, intestinal barrier function, and immunity (9,10). Conversely, insoluble fibers, such as cellulose, are less fermentable (1,4), yet contribute to laxation.

In addition to fiber, dietary fat can influence gastrointestinal and metabolic health (11) through primary bile acid synthesis (12), and ileal reabsorption, with subsequent production of pro-inflammatory secondary bile acids such as deoxycholic (DCA) and lithocholic (LCA) acid, by gut microbes (8). High-fat diets (> 35% of total kcal), particularly high in saturated fat and low in fiber, increase secondary bile acids, which are associated with gallstone formation and gastrointestinal cancers (8). In contrast, unsaturated fats such as MUFAs are linked to favorable metabolic and inflammatory profiles (13), potentially influencing bile acid metabolism and gastrointestinal health.

Although fiber and unsaturated fats influence the gut microbiome and overall health, individuals consume whole foods as part of dietary patterns. Whole foods contain nutrients, phytochemicals, and other bioactive compounds that are packed and compartmentalized within a food matrix that cannot be entirely replicated by isolated nutrients (11). As such, whole foods within dietary patterns provide additional compounds and nutrients that influence microbial metabolism (14) and physiological outcomes (11). Thus, foods that are naturally rich in fiber and unsaturated fats, such as avocados, provide a unique opportunity to examine how the food matrix modulates gastrointestinal and metabolic health as well as the gut microbiome.

Avocado consumption has increased significantly over the past decade (15) and has been shown to reduce cardiovascular disease risk, support weight management, and improve cognitive function (16). Avocado’s nutrient profile, including fiber (pectin, xyloglucans, cellulose) and oleic acid (MUFA), as well as polyphenols, phytosterols, carotenoids, vitamins, and minerals, likely contributes to these benefits (13,16,17). Previous research from our laboratory showed that daily avocado consumption increases gut microbiota alpha diversity, fecal acetate concentrations, and *Faecalibacterium* abundance, while reducing secondary bile acids (i.e., LCA) in adults with overweight and obesity (18).

Currently, there is limited understanding of how the combination of fibers, fat, and other compounds in whole foods (i.e., avocados) affects physiological outcomes. Clinical research has historically focused on isolated fibers or specific dietary patterns without examining specific foods or food matrices (19). Thus, to address this gap and extend our previous work, we aim to investigate how consuming whole fresh Hass avocados - beyond their fiber and MUFA content - affects gastrointestinal health and microbiome in adults with overweight and obesity. We hypothesized that the combination of fiber and MUFA in avocados, as compared to control groups, would support gastrointestinal health by (1) increasing *Faecalibacterium spp.* and SCFA concentrations, while (2) decreasing DCA and LCA-producing bacteria and bile-salt hydrolase genes, which would be proportional to the reductions in DCA and LCA concentrations as well as circulating LBP.

## METHODS

The procedures for this study were conducted in accordance with the ethical standards outlined in the Declaration of Helsinki and were approved by the Office for the Protection of Research Subjects at the University of Illinois, Urbana-Champaign (IRB protocol #22788). All study participants provided written informed consent.

### Participants

A total of 57 adults without diabetes, aged 25-74 years, with overweight and obesity (BMI ≥ 25kg/m^²^), were randomized to the study. All participants were informed of the study’s purpose and experimental procedures during phone screening prior to providing written consent. The exclusion criteria of the study included avocado and food allergies or intolerance, prior diagnosis of liver or gastrointestinal disease, elevated fasting blood glucose (>126mg/dl), elevated blood pressure (>160/100mmHg), pregnancy/breastfeeding, use of tobacco, alcohol consumption (> 2 beverages/day), oral antibiotics during the previous 6 weeks, lipid-lowering medications, oral hypoglycemics or insulin, history of malabsorptive or bariatric surgeries, allergy to latex and concurrent enrollment in another dietary, exercise or medication study, and inability to consume study meals. Participants underwent a baseline visit while fasting to confirm eligibility criteria. At baseline, measurements of weight and height were taken to verify BMI, blood pressure was recorded, and fasting blood glucose and liver enzymes [including alanine aminotransferase (ALT), aspartate aminotransferase (AST) and alkaline phosphatase (ALP)] were quantified.

### Study design

The *Persea americana* for total health 2 (PATH-2) study was a randomized, controlled, counterbalanced, crossover, complete feeding trial (NCT05941728) conducted between July 2023 and August 2024 at the University of Illinois Urbana-Champaign (UIUC). Each participant underwent three 4-week dietary periods: avocado (AV), Average American dietary control (AA), and oleic acid + fiber nutrients (OF), separated by a 2-week washout period. Participants were randomly assigned to one of six possible treatment sequences: ABC/ACB/BAC/BCA/CAB/CBA (**Supplmental Figure 1**). The random allocation sequence was generated using a computer-based random number generator in Microsoft Excel. Research staff remained blinded for the duration of the study. The base diet of all three conditions consisted of a 7-day cycle menu containing typical American foods with an Acceptable Macronutrient Distribution Range (AMDR) of 45% CHO, 35% fat, and 20% protein. Meal preparation occurred within our on-site metabolic kitchen, with each ingredient weighed to the nearest gram by study staff. Participants’ energy requirements were scaled as needed during meal pick up twice a week to ensure weight maintenance during each dietary period. The study condition base diets were isocaloric. However, compared to the AA group, the AV and OF conditions provided greater amounts of MUFA and fiber (**Table 1**).

**Figure 1.**
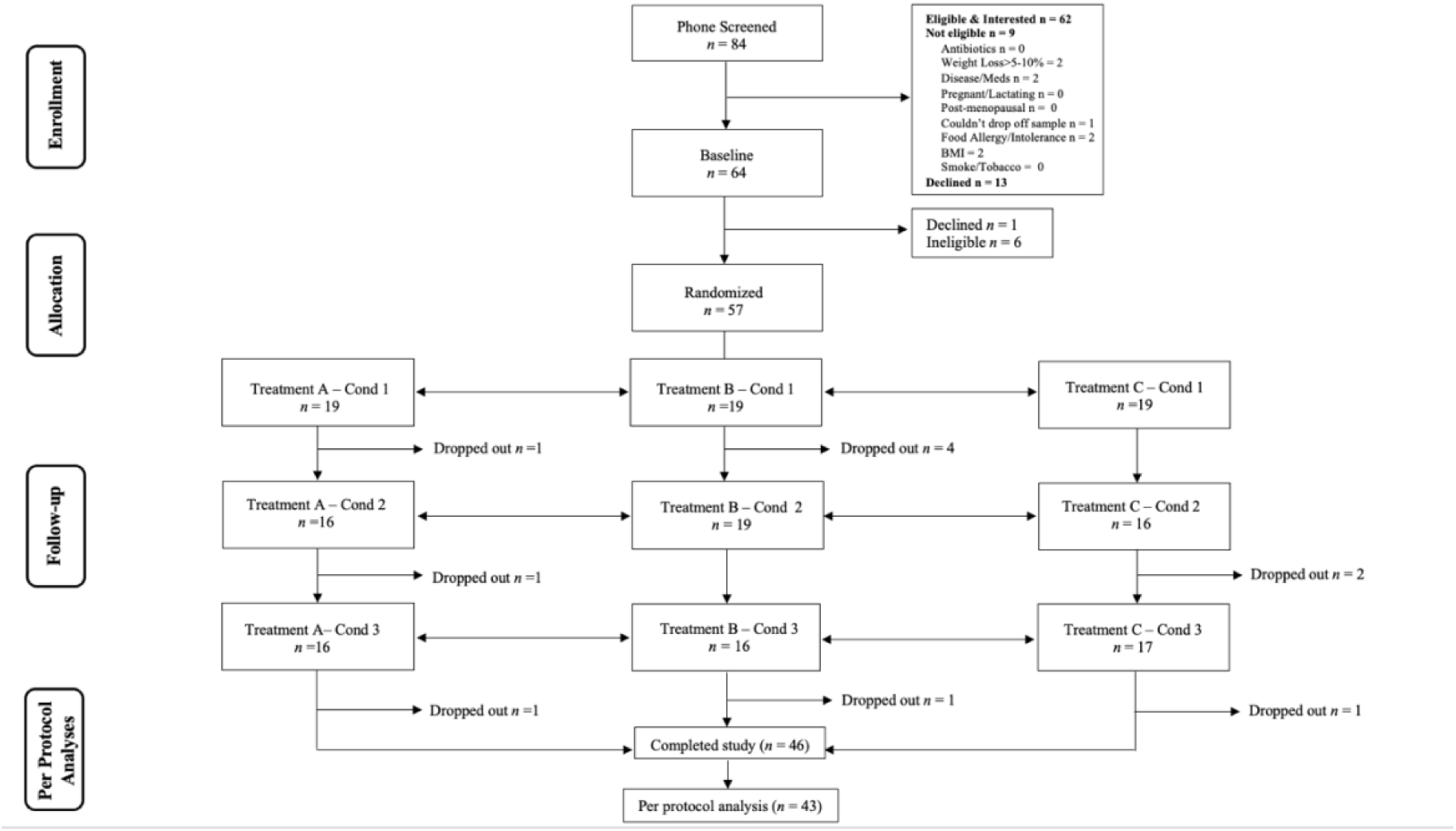
CONSORT flow diagram of recruitment and randomization. Each “n” indicates the number of participants at each stage of the study. Participants were randomized to avocado (A), nutrients (B) or control (C)

**Table 1.** Dietary Condition Nutrient Information.

| Nutrient | Control (AA) | Nutrients (OF) | Avocado (AV) |
| --- | --- | --- | --- |
| Calories, kcal | 2,000 | 2,003 | 2,003 |
| CHO, % | 45.7 | 46.2 | 46.6 |
| Fiber, g | 14.8 | 22.8 | 23.1 |
| Total Fat, % | 35.3 | 34.2 | 36.0 |
| SFA, g | 30.2 | 20.7 | 21.3 |
| PUFA, g | 16.1 | 13.4 | 12.7 |
| MUFA, g | 17.9 | 28.7 | 31.0 |
| Protein, % | 18.8 | 18.4 | 18.4 |
| SFA = Saturated fatty acid; PUFA = Polyunsaturated fatty acids;<br>MUFA = Monounsaturated fatty acids |  |  |  |

In the AV condition, participants consumed 209g of avocado at dinner. Avocados were sourced from the Hass Avocado Board and processed in the metabolic kitchen. Upon arrival, the avocados were left to ripen at room temperature (15-20°C) for up to 5 days. The avocados were peeled and cut to weigh 209g and then vacuum sealed to prevent browning. Participants were instructed to consume the avocado as provided, without mixing it or altering its food matrix. Each serving of avocados contained approximately 21g of MUFA, 5g of saturated fat and 9g of fiber, of which 5g (56%) and 4g (44%) are soluble and insoluble fiber, respectively (**Table 2**). The OF group consumed a vanilla pudding/pie crust recipe that mimicked the macronutrient and caloric profile of avocados. For this pudding, low-methoxy (LM) homogalacturonan (HG) pectin was sourced from CP Kelco, and cellulose was purchased from *NutriCology* as microcrystalline cellulose. Overall, one serving of the OF pudding/pie crust recipe contained 18g of MUFA, 4g of SFA, and 8g of dietary fiber, of which 3g (37.5%) and 5g (62.5%) are soluble and insoluble fiber, respectively (Table 2). The AA group consumed a similar vanilla pudding, with the same macronutrient and calorie content as the avocado serving; however, the pie crust recipe was lower in MUFA (9g) and fiber (2g), and higher in saturated fat (12g) (Table 2).

**Table 2.** Comparison of nutrients from avocado relative to control snacks.

| Nutrient | Control (AA) | Nutrients (OF) | Avocado (AV) |
| --- | --- | --- | --- |
| Portion size, g | 94 | 91 | 209 |
| Calories, kcal | 349 | 349 | 349 |
| CHO, g | 20 | 16 | 18 |
| Fiber, g | 2 | 8 | 9 |
| Soluble, g | 0.5 | 5 | 5 |
| Insoluble, g | 1.5 | 3 | 4 |
| Total Fat, g | 28 | 28 | 32 |
| SFA, g | 12 | 4 | 5 |
| PUFA, g | 3 | 5 | 4 |
| MUFA, g | 9 | 18 | 21 |
| Protein, g | 4 | 4 | 4 |
| SFA = Saturated fatty acid; PUFA = Polyunsaturated fatty acids; MUFA = Monounsaturated fatty acids |  |  |  |

### Experimental protocol

At the end of each 4-week dietary intervention, participants provided blood, fecal, and urine samples. Participants provided a total of two fecal samples between days 22-26 of each dietary condition. Between days 25-28, participants underwent a study visit following a 10-hour fast that included anthropometrics, blood draw and intestinal permeability test (Supplemental Figure 1).

### Anthropometrics

Weight (kilograms) and height (centimeters) were measured in triplicate using a Seca model 769 electronic scale and a wall-mounted stadiometer, respectively. BMI was calculated as kg/m^2^. Waist, umbilicus, and hip circumferences (all centimeters) were measured using a measuring tape. Blood pressure and pulse were assessed using an automated Omron oscillometric device. Participants were instructed to stay seated for five minutes prior to blood pressure measurement with feet flat on the floor, back supported, and the appropriately sized cuff placed on the upper arm at heart level. All measurements were done in triplicate, with blood pressure measurements taken at least 2 minutes apart. Results are presented as averages of all three measurements across each dietary condition.

### Fecal collection and processing

At the end of each dietary condition (days 22-26), participants were asked to provide two fecal samples. Participants were instructed to provide fecal samples within 15 minutes of defecation using provided materials, including Commode Specimen Collection Systems (Sage Products), ice packs, and coolers. A trained research team member received the sample and processed it. The fecal samples were homogenized upon arrival in the laboratory, aliquoted into cryovials, flash-frozen in liquid nitrogen, and stored at -80°C until further analysis. A pH measurement (Denver Instrument) and the consistency of the fecal sample according to the Bristol Stool Chart (BSC) were recorded for each sample. Fecal samples were also aliquoted into 15 ml test tubes for phenol and indole quantification, pre-weighed aluminum tins to for dry matter determinations, and acidified with 2N-HCL (10%wt: vol) for SCFAs (acetate, propionate, butyrate), BCFAs (valerate, isovalerate, isobutyrate), and ammonia analyses. These aliquots were all frozen at -20°C until further analysis.

### Fecal metabolites

Fecal dry matter was determined according to the Association of Analytical Chemists, 1984 (20), using fresh fecal samples to allow for reporting of fecal fermentative end products on a dry matter basis (DMB). Ammonia concentrations were measured according to Chaney and Marbach (21). The sample aliquots for SCFA and BCFA were thawed at room temperature for 15 min and centrifuged at 13,000g for 10 min. Samples were analyzed through gas chromatography mass spectrometry (GC-MS), normalized by dry weight. Phenols and indoles were extracted from fecal samples as previously described (22). Bile acids were assessed using liquid chromatography-mass spectrometry (LC-MS) by the Carver Metabolomics Core of the Roy J. Carver Biotechnology Center, University of Illinois Urbana-Champaign, using a targeted approach. Briefly, unconjugated primary (cholic acid [CA] and chenodeoxycholic acid [CDCA]) and secondary (deoxycholic acid [DCA], lithocholic acid [LCA], and ursodeoxycholic acid [UDCA]) as well as the glycine and taurine conjugated forms were quantified. Peak integration and quantitation were done using calibration curves adjusted for internal standards with MultiQuant v3.1 software (Sciex, Framingham, MA, USA). Data imputation was performed on CA, DCA, TCDCA, and TDCA, with 94%, 99%, 88%, and 89% of data available, respectively, using the “purrr” (v1.1.0) and “tibble” (v3.3.0) packages. Specifically, instances where the value was below the detectable limit, values were imputed by calculating half the minimum value for the respective bile acid. For each participant and condition, SCFA, BCFA, phenol, indole, and bile acid concentrations were calculated as the average concentration of two fecal samples (n = 258).

### Gut microbiome composition and function

Fecal microbial DNA was extracted using the [DNeasy PowerLyzer PowerSoil Kit](https://www.qiagen.com/us/products/discovery-and-translational-research/dna-rna-purification/dna-purification/microbial-dna/dneasy-powerlyzer-powersoil-kit) (Qiagen, Germantown, MD, USA). At the end of each DNA extraction, the quantity of DNA was measured using a Qubit 3.0 Fluorometer. DNA quality was assessed via gel electrophoresis on a 1% agarose gel. After the quality assurance, the two fecal samples from each condition were pooled in equimolar amounts for sequencing. The metagenome for all 129 samples was sequenced at the Roy J. Carver Biotechnology Center DNA Services Unit at the University of Illinois, Urbana-Champaign. The gDNA libraries were sequenced on a NovaSeq X Plus on a 25B lane with 2x150nt reads and produced over six billion reads with great quality scores (Phred score ≥ 30).

Data processing and bioinformatics were conducted using the Carl R. Woese Institute for Genomic Biology (IGB) Biocluster at the University of Illinois Urbana-Champaign.

Firstly, pre-processing was performed on the paired-end FASTQ input files using KneadData (v0.12.0) to remove low-quality reads and adapters (via Trimmomatic) as well as host DNA (using the KneadData database 20230405). Tandem Repeat Finder (TRF), as part of KneadData, was used to identify and remove repetitive DNA sequences. FastQC was used to evaluate the quality of the reads before and after processing. Lastly, the R1/R2 cleaned files were concatenated into one file for downstream analysis.

Metagenomic Phylogenetic Analysis 4 (MetaPhlAn 4) (v4.0.6) was used to perform taxonomic profiling, which used Bowtie2 to align reads to the ChocoPhlAnSGB_202212 database. The kneaddata-filtered FASTQ files were analyzed using -t rel_ab_w_read_stats to measure relative abundance, which normalizes taxonomic abundance while preserving read counts. Afterwards, the samples were merged to generate study-wide abundance tables. The *merge_metaphlan_tables.py* script was used to create a table with the percentage of relative abundance of taxa (rows) across all samples (columns). A modification to the previous script, *merge_metaphlan_tables_abs.py,* was made to estimate the absolute read counts for statistical analysis. The script requires MetaPhlAn to be run with -t rel_ab_w_read_stats to capture both relative abundance and observed counts (23).

The HMP Unified Metabolic Analysis Network (HUMAnN v3.7) was used to assess the functional capacity of the microbial communities in our dataset, using the taxonomic profiles generated by MetaPhlAn. The analysis used a nucleotide database (ChocoPhlAn v201901b) and a protein database (UniRef90 v201901b) to stratify gene functions at the species level. Similarly, the samples were merged to obtain gene families, pathway abundance, and pathway abundance information. The tables were normalized to relative abundance using “total sum scaling” normalization, where each sample is constrained to sum to 1. The *human_rename_table* script from HUMAnN was used to rename IDs from UniRef90 to readable annotations for downstream analysis. To further simplify the exploration of gene family abundance data, we regrouped gene families into KEGG Orthology (KO) using *humann_regroup_table*. Lastly, we used the *humann_split_stratified_table* script to split the data into stratified and unstratified gene families by species.

We selected the SCFA and bile acid metabolism-related pathways and KEGG orthologs to align with our initial research aims and also to limit the number of KOs investigated. The mapping files were imported using R-package KEGGREST (v1.48.1) to create a filtering vector that included “Primary bile acid biosynthesis” (ko00120), “Secondary bile acid biosynthesis” (ko00121), “Pyruvate metabolism” (ko00620), “Propionate metabolism” (ko00640), “Butanoate metabolism” (ko00650), “Carbon metabolism” (ko00640), “Glyoxylate and dicarboxylate metabolism” (ko00640) “Glycolysis/Gluconeogenesis” (ko00010), “Pentose phosphate pathway” (ko00030), “Fructose and mannose metabolism” (ko00051) and “Starch and sucrose metabolism” (ko00500).

### Differential microbial and gene abundance analysis

Differential abundance analysis was performed with the Microbiome Multivariable Associations with Linear Models 3 (MaAsLin3) package in R (24), using the 100 most abundant taxa and species. To ensure robustness, we applied a minimum threshold of 10 reads per sample and a prevalence of at least 10%, while excluding unclassified taxa (^GGB) and species (^s GGB) from MetaPhlan. After these filtering steps, 67 genera and 86 species remained for differential abundance analysis. Briefly, raw counts were normalized by total sum scaling (TSS) to obtain relative abundances, and per-sample library sizes were provided within the *unscaled_abundance* parameter in the maaslin3() function. The procedure rescales relative abundances to estimate absolute abundance for each taxon according to the formula:

𝐴𝑏𝑠𝑜𝑙𝑢𝑡𝑒 𝑎𝑏𝑢𝑛𝑑𝑎𝑛𝑐𝑒_𝑖𝑗_ = 𝑅𝑒𝑙𝑎𝑡𝑖𝑣𝑒 𝑎𝑏𝑢𝑛𝑑𝑎𝑛𝑐𝑒_𝑖𝑗_ 𝑥 𝑇𝑜𝑡𝑎𝑙 𝑅𝑒𝑎𝑑𝑠_𝑖_.

Absolute abundances were log-transformed, and linear mixed-effects models were fit with treatment as a fixed effect and participant as a random effect. Median compositionality correction was disabled because the total abundance normalization accounts for compositionality of the data (24). For gene abundance, we filtered based on pathways related to bile acid synthesis and SCFA, as previously described. Then, we calculated the mean prevalence and abundance of gene families from the related pathways using the summary function to establish parameters that stabilize the models. For bile acids, the mean prevalence and abundance of gene families were 1% and 0.00001%, respectively, resulting in the selection of 16 of the 31 annotated genes. For SCFA and carbohydrate metabolism enzymes, the mean prevalence and abundance of genes were 5% and 0.00001%, respectively, thereby selecting 1,372 of the 3,065 and 1138 of the 2314 gene families for SCFA and carbohydrate metabolism, respectively, for statistical analysis. These filtered data sets were fed into the maaslin3() function to assess differential gene abundance across dietary conditions.

### Carbohydrate-active enzymes (CAZyme) annotation

Carbohydrate-active enzyme (CAZyme) profiles were obtained from UniRef90 functional annotations stratified by species. The CAZyme mapping database, available at [GitHub](https://github.com/junhuili/cazy_uniref90_humann/blob/main/map_cazy_uniref90.txt), was used to filter UniRef90 gene families obtained by HUMAnN. Similar to SCFA and bile acid gene abundance, we calculated the mean prevalence and abundance of CAZymes. Of the 1921 CAZymes in our data set, a total of 905 CAZymes were found in 5% of the samples, with a mean abundance of 0.00001%. This filtered data set was fed into the maaslin3() function to assess differential gene abundance across dietary conditions.

### Alpha and beta diversity analysis

Alpha and beta diversity were performed in R with the “vegan” package (v2.7.1) using the taxonomic files retrieved from MetaPhlAn. The relative abundance of taxa at the genus level was used to calculate and assess alpha diversity. The specnumber() and the diversity() function were used to calculate richness and the Shannon index, respectively. For beta diversity, we used the genus relative abundance files retrieved from MetaPhlAn. Briefly, the cmdscale() function was used to calculate Principal Coordinate Analysis (PCoA) with Bray-Curtis distance metric, calculated by vegdist(). Then, we conducted a PERMANOVA analysis using the adonis2() function with 5000 permutations to assess the effect of treatment on community composition. Inside the adonis2() function, we added a strata component to account for repeated measures design. To assess data dispersion between treatments, the betadisper() function was used with the Bray-Curtis distance method calculated.

### Correlation and network analysis

Co-occurrence network analysis, including correlation analysis, was performed using the R package “Hmisc” (v5.2.4) (25). For the network analysis, we utilized the filtered data frame obtained from the differential species abundance as well as significant host and microbial phenotypic data, including fecal and urine metabolites, inflammatory markers, and anthropometrics in both AV and OF conditions. Then, we visualized the network and clusters of microbial species using a heatmap developed by the R package “ComplexHeatmap” (v2.24.1) and “circlize” (0.4.16) (25). Using Spearman’s rank correlation, the correlation analysis was performed with pooled data from all dietary conditions. The p-values were adjusted for multiple comparisons using the Benjamini-Hochberg false discovery rate (FDR) method, with significance reported at α ≤ 0.10. This was an exploratory analysis assessing the association between microbial species and multi-omics variables.

### Blood assessment

Blood was taken at the 4-wk appointment visit following an overnight fast (≥10-hr). Blood samples were collected in serum, heparin, and EDTA tubes. EDTA plasma samples were centrifuged at 1,300g for 10min and stored at -80°°C until further analysis of lipopolysaccharide binding protein (LBP), tumor necrosis factor α (TNF-α), interleukin-6 (IL-6), and C-reactive protein (CRP). Systemic inflammatory markers were assessed using the colorimetric enzyme-linked immunosorbent assay (ELISA) technique according to the manufacturer’s instructions. LBP was assessed using the Hycult Biotech (HK315); TNF-α, IL-6 and CRP was assessed using the R&D systems kit HSTA00E, HS600C, and DCRP00B, respectively.

### Intestinal inflammation

The quantitative determination of secretory immunoglobulin A (sIgA) and calprotectin was measured using the 30-SECHU-E01 and 30-CALPHU-E01 ELISA kits from ALPCO Diagnostics, respectively. Fecal samples were thawed and homogenized using an inoculation loop. The extraction of sIgA and calprotectin was done using the manual extraction method. Briefly, thawed fecal samples (15-20mg) were diluted (1:100) with extraction buffer, vortexed for 30 min, and the supernatant (40 μL) was used for further dilutions and downstream analysis as established by each protocol.

### Intestinal permeability

Intestinal permeability was assessed by measuring the urinary excretion of monosaccharides (rhamnose and mannitol) and disaccharides (sucralose). At the appointment visit at the end of each study condition (and after the fasted blood draw), participants consumed a beverage with 0.5g mannitol, 0.2g rhamnose, and 1g sucralose mixed with 155ml of water. All urine produced was collected for 24 hours and divided at different timepoints (T) following consumption: 0-2 hrs (T1), 2-8 hrs (T2), and 8-24 hrs (T3). The urine aliquoted between 0-2 hrs after beverage consumption reflects small intestine permeability, whereas the urine collected between 8-24 hrs reflects colonic permeability (26,27). The sugars in urine were measured through gas chromatography. Urinary sugar was quantified through GC at the Carver Metabolomics Core of the Roy J. Carver Biotechnology Center, University of Illinois Urbana-Champaign. The data from GC (μg/ml) was used to calculate urinary sugar recovery (%) using the following formula:

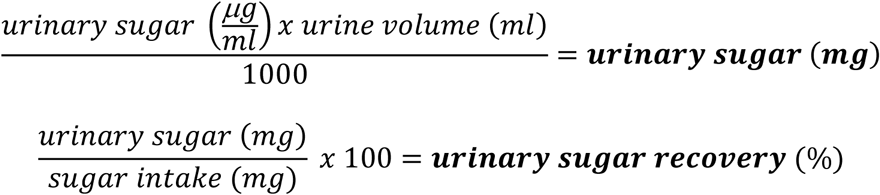

### Gastrointestinal tolerance, regularity, and stool consistency

Gastrointestinal symptoms, bowel movements, and stool characteristics were measured using a self-reported record of the last 7 days of each treatment. The 7-day self-reported record included subjective ratings 1 (absent), 2 (mild), 3 (moderate), and 4 (severe) for the following gastrointestinal symptoms: burping, cramping/pain, distension/bloating, flatulence/gas, nausea, reflux (heartburn), and rumblings. Participants also recorded their bowel movement consistency using the Bristol Stool Scale: (1 [separate hard lumps], 2 [sausage-shaped but lumpy], 3 [sausage-like but with cracks on the surface], 4 [sausage-like, smooth, and soft], 5 [soft blobs with clear cut edges], 6 [fluffy pieces, mushy], and 7 [entirely liquid]) and ease of passage (1 [very easy], 2 [easy], 3 [neither easy nor difficult], 4 [difficult], 5 [very difficult]).

## STATISTICAL ANALYSIS

Participant baseline characteristics are presented as means ± SDs for continuous variables, and sample size (n) with percentages for categorical variables. The primary outcomes of the study, including differential abundance of *Faecalibacterium spp.*, SCFA and secondary bile acid concentrations, abundance of bile salt hydrolase genes, and circulating LBP, were analyzed using R (v4.5.1). All variables are reported with 95% confidence intervals and standard errors (SE) in the tables. Using both *Faecalibacterium* and acetate results from our previous trial (18), our *a priori* power calculation indicated a sample size of 39 [moderate effect size (Cohen’s d = 0.39), 1-sided α of 0.10, and 80% power]. To account for attrition, 57 participants were enrolled.

To conduct the analysis of variance (ANOVA), normality of residuals and homogeneity of variance were checked using the Shapiro-Wilk normality test (*W* ≥ 0.9) and Levene’s test, respectively, with the “car” (v3.1.3) package. To address non-normality and heteroscedasticity, we applied the Box–Cox power transformation to the predictor variables (28) using the boxcox() function from the “MASS” (v7.3.65) package in R. The optimal lambda (λ) was estimated by selecting the value that maximized the log-likelihood within the range −2 to 2. Variables were log-or square root transformed as per the lambda parameter to meet normality assumptions. A non-parametric Friedman test for repeated measures was used when transformations failed to meet the assumptions of ANOVA.

Differential abundance analysis at the genus and species levels was conducted using the Microbiome Multivariable Associations with Linear Models 3 (MaAsLin3) package (v1.0.0) in R (24). Following the filtration and quality control steps described previously, we fitted a generalized mixed effects model with treatment as a fixed effect and participant as a random effect (24) adjusted with Benjamini-Hochberg false discovery rate (FDR) corrections. Similarly, gene functional capacity stratified by species was analyzed using MaAsLin3. The annotated KEGG tables generated by HUMAnN were normalized using the *human_renorm_table* function and log-transformed to stabilize variance. The results for microbial and gene abundance are expressed using β-coefficients, which indicate the unit change in abundance within AV and OF, compared to AA. Significance level for microbial abundance was defined at FDR q ≤0.15 and gene abundance at FDR q≤0.25. Lastly, statistical analysis for microbial diversity was assessed using the non-parametric Friedman test for α-diversity and PERMANOVA with 5000 permutations for β-diversity.

Fecal metabolites, including SCFAs, BCFAs, phenols, indoles, ammonia, and bile acids, were analyzed employing linear mixed models (LMMs) using “lme4” (v1.1.37) and “lmerTest” (v3.1.3) R packages, with treatment as a fixed effect and participant as a random effect. Period (diet order) was tested as a fixed effect for all outcomes to assess potential carryover effects; since it did not significantly contribute to variance, it was excluded from the final models. *Post hoc* pairwise contrasts were estimated applying least-square means using the “emmeans”( v1.11.2.8) package and adjusted with Benjamini-Hochberg corrections. The markers of barrier function [urinary excretion (mg) and recovery (%)], as well as LBP, were evaluated using the non-parametric Friedman test. The remaining variables assessed, such as pH, IL-6, TNF-α, CRP, fecal calprotectin, sIgA flatulence, BSC, stool ease, number of bowel movements, weight, blood pressure, body weight, waist circumference, and umbilicus were analyzed using LMMs, while gastrointestinal symptoms, including rumbling, burping, bloating, cramping, nausea, and reflux were assessed using non-parametric Friedman test. The significance level for fecal metabolites, anthropometric measurements, gastrointestinal symptoms, and inflammatory markers was defined as α ≤ 0.05.

For variables that were significant (α ≤ 0.05) or trending toward significance (α ≤ 0.10), effect sizes were estimated using the model_parameters function from the “parameters” (v0.28.2) package in R. Linear regression models were refit using standardized z-scores, and confidence intervals were computed using the Satterthwaite method for mixed effects models. Beyond statistical significance, the coefficients enabled us to quantify and compare the magnitude and direction of the treatment effect across significant variables. For data visualization purposes only, data points from total primary bile acids, CA species, and CDCA species were winsorized at the 95^th^ percentile, with bar plots and standard error using ordinal means.

## RESULTS

### Participant characteristics

Baseline demographic and clinical characteristics are presented in **Table 3** and **Supplemental Table 1**, respectively. The study consort form (**Figure 1**) details the study screening, recruitment, and retention across dietary conditions. Of the 57 participants randomized, 46 completed the study, resulting in an attrition rate of 19% (n = 11). Dropouts from the study are due to scheduling conflicts (n = 6), family health issues (n = 1), and difficulty adhering to study diets (n = 4). Furthermore, among the participants who completed the study, forty-three (n = 43) were included in the per-protocol data analysis based on ≥ 70% dietary compliance in all three conditions, without the addition of other foods/beverages as measured by their compliance logs and twice-weekly interviews with study staff.

**Table 3.** Baseline Demographic of Participants Randomized to Control (AA), Nutrients (OF) and Avocado (AV)

|  | Overall<br>(n=43) | Control<br>(AA) (n=16) | Nutrients<br>(OF) (n=12) | Avocado (AV)<br>(n=15) |
| --- | --- | --- | --- | --- |
| **Education** |  |  |  |  |
| Some College | 5 (11.6%) | 2 (12.5%) | 2 (16.7%) | 1 (6.7%) |
| University Graduate | 11 (25.6%) | 4 (25%) | 5 (41.7%) | 2 (13.3%) |
| Master's Degree | 22 (51.2%) | 8 (50%) | 4 (33.3%) | 10 (66.7%) |
| PhD or equivalent | 5 (11.6%) | 2 (12.5%) | 1 (8.3%) | 2 (13.3%) |
| **Race** |  |  |  |  |
| Asian | 6 (13.9%) | 2 (12.5%) | 0 (0%) | 4 (26.7%) |
| Black or African American | 4 (9.3%) | 0 (0%) | 3 (25%) | 1 (6.7%) |
| Mixed or Other | 5 (11.6%) | 1 (6.2%) | 4 (33.3%) | 0 (0%) |
| White or Caucasian | 28 (65.1%) | 13 (81.2%) | 5 (41.7%) | 10 (66.7%) |
| Latino | 3 (6.9%) | 0 (0%) | 3 (25%) | 0 (0%) |
| **Sex** |  |  |  |  |
| Female | 24 (55.8%) | 11 (68.8%) | 5 (41.7%) | 8 (53.3%) |
| Male | 19 (44.2%) | 5 (31.2%) | 7 (58.3%) | 7 (46.7%) |
| **Income** |  |  |  |  |
| \$10,000-20,000 | 1 (2.32%) | 0 (0%) | 1 (8.3%) | 0 (0%) |
| \$21,000-30,000 | 1 (2.32%) | 0 (0%) | 1 (8.3%) | 0 (0%) |
| \$31,000-40,000 | 4 (9.3%) | 3 (18.8%) | 1 (8.3%) | 0 (0%) |
| \$41,000-50,000 | 5 (11.6%) | 1 (6.2%) | 1 (8.3%) | 3 (20%) |
| \$51,000-60,000 | 4 (9.3%) | 1 (6.2%) | 3 (25%) | 0 (0%) |
| \$61,000-70,000 | 3 (6.9%) | 3 (18.8%) | 0 (0%) | 0 (0%) |
| \$71,000-80,000 | 4 (9.3%) | 1 (6.2%) | 1 (8.3%) | 2 (13.3%) |
| \$81,000-90,000 | 2 (4.6%) | 1 (6.2%) | 0 (0%) | 1 (6.7%) |
| \$91,000-100,000 | 4 (9.3%) | 3 (18.8%) | 0 (0%) | 1 (6.7%) |
| Greater than \$100,000 | 15 (34.9%) | 3 (18.8%) | 4 (33.3%) | 8 (53.3%) |
| **Mode of Birth** |  |  |  |  |
| Cesarean section birth | 6 (13.9%) | 4 (25%) | 2 (16.7%) | 0 (0%) |
| Vaginal birth | 35 (81.4%) | 11 (68.8%) | 10 (83.3%) | 14 (93.3%) |
| Not known | 2 (4.6%) | 1 (6.2%) | 0 (0%) | 1 (6.7%) |
| **Breastfeeding status** |  |  |  |  |
| Yes | 23 (53.5%) | 10 (62.5%) | 4 (33.3%) | 9 (60%) |
| No | 12 (27.9%) | 4 (25%) | 5 (41.7%) | 3 (20%) |
| Unknown | 8 (18.6%) | 2 (12.5%) | 3 (25%) | 3 (20%) |
| **Age (years)** | 41.7 ± 9.73 | 41.2 ± 8.11 | 41.8 ± 13.5 | 42.1 ± 8.38 |

### Anthropometrics

For the analysis that follows, one participant during the AA and AV conditions, and another participant during the OF condition, were unable to attend study visits due to an unrelated illness. Further, due to procedural oversight, weight and anthropometric circumference measurements were not collected for one participant in the OF condition. Lastly, blood pressure measurement could not be taken for one participant in the AA group due to equipment failure. This resulted in different sample sizes across dietary conditions for weight, anthropometric circumference, and blood pressure measurements, as depicted in **Table 4**. Weight, natural waist circumference, and umbilical waist circumference, as well as systolic blood pressure and pulse, remained unchanged across dietary conditions (all p > 0.32). Notably, there was a mean decrease in diastolic blood pressure of 2.3 mmHg and 0.8 mmHg in the AV and OF groups, respectively, compared to the AA group, which was marginally significant (p = 0.052) (Table 4). Furthermore, the lower diastolic blood pressure within the AV group suggests a moderate effect size in this treatment group (β = - 0.24, p = 0.01) (**Supplemental Table 2**).

**Table 4.** Anthropometrics, blood pressure, and pulse rate across dietary conditions.

|  | Control (AA) |  | Nutrients (OF) |  | Avocado (AV) |  | P value |
| --- | --- | --- | --- | --- | --- | --- | --- |
|  | Mean (95% CI) [n] | SEM | Mean (95% CI) [n] | SEM | Mean (95% CI) [n] | SEM |  |
| Weight, kg | 95.3 (88.8-102) [42] | 3.19 | 95.0 (88.6-101) [41] | 3.2 | 95.1 (88.7-102) [42] | 3.19 | 0.76 |
| Natural WC, cm<sup>1</sup> | 96.6 (92.7-101) [42] | 1.99 | 97.1 (93.2-101) [41] | 2 | 96.3 (92.4-100) [42] | 1.98 | 0.39 |
| Umbilicus WC, cm | 104 (98.9-109) [42] | 2.48 | 103 (98.4-108) [41] | 2.48 | 104 (99.0-109) [42] | 2.48 | 0.61 |
| Systolic, mmHg | 121 (117-125) [41] | 2.03 | 121 (117-125) [42] | 2.02 | 119 (115-123) [42] | 2.02 | 0.32 |
| Diastolic, mmHg | 82.9 (79.9-86.0) [41] | 1.52 | 82.1 (79.1-85.2) [42] | 1.51 | 80.6 (77.5-83.6) [42] | 1.52 | 0.052 |
| Pulse | 68.6 (65.8-71.4) [41] | 1.4 | 70.1 (67.3-72.8) [42] | 1.39 | 69.6 (66.8-72.4) [42] | 1.39 | 0.36 |

Data are presented as estimated marginal means (EMMs) along with 95% confidence intervals (CI) in parenthesis and sample size [n] in brackets. P-values are derived from linear mixed models (LMMs) with treatment as a fixed effect and participant as the random effect.
<sup>1</sup>Variable was log transformed to meet normality and homogeneity of variance assumptions for repeated measures ANOVA. WC = waist circumference.

### Fecal metabolites

For this analysis, five participants did not provide a second fecal sample; therefore, only one sample was used for analysis for those 5 individuals (n = 253). For SCFA, BCFA, phenol, and indole analyses, two participants did not provide sufficient sample volume to measure these metabolites (n = 251). For bile acids, an additional ten participants had insufficient samples after processing for SCFA, and DNA extraction (n = 241).

Both AV and OF group showed higher acetate levels by 68 µmol/g DMB (28%) and 44 µmol/g DMB (18%), respectively, compared to the AA group (p < 0.001 & p = 0.019) (**Supplemental Table 3; Figure 2A**). Similarly, AV and OF groups exhibited higher total SCFA concentrations by 72 µmol/g DMB (19%) and 58 µmol/g DMB (15%), respectively, compared to AA (p = 0.018 & p = 0.037), primarily driven by acetate. To assess the effect and magnitude of the relationship between acetate, total SCFA, and treatment, we measured the standardized β coefficients of the linear regression model. Both AV and OF had positive associations with acetate (β = 0.51 [p < 0.001]; β = 0.32 [0.01]), and total SCFA (β = 0.29 [p = 0.02]; β = 0.36 [p < 0.01), respectively (Supplemental Table 2).

**Figure 2.**
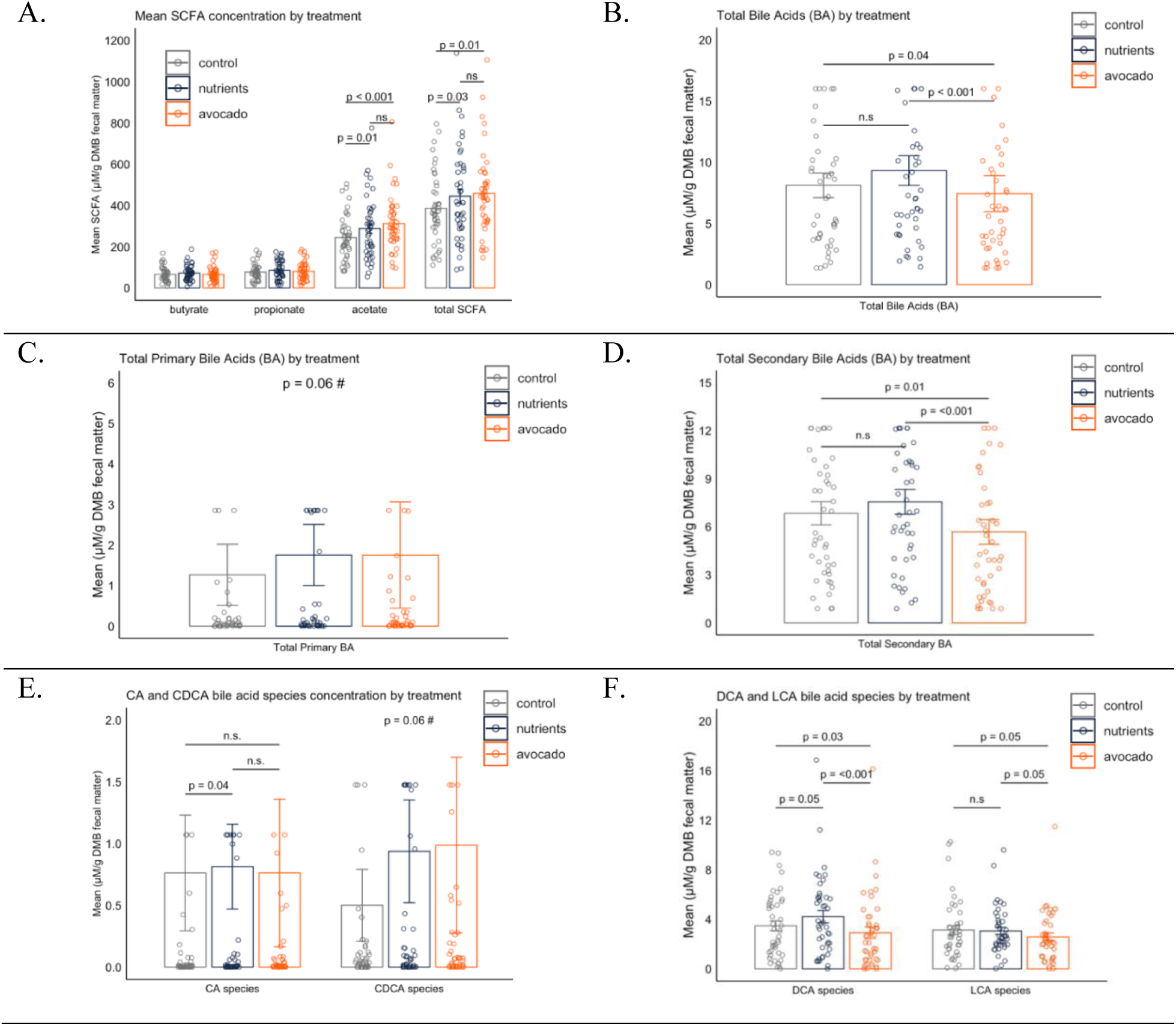
Mean SCFA and bile acid concentrations across dietary conditions. Overview of fecal SCFA and bile acids across dietary conditions with (A) mean acetate, propionate, butyrate and total SCFAs, (B) total bile acids, (C) total primary bile acids, (D) total secondary bile acids, (E) cholic (CA) and chenodeoxycholic (CDCA) acid species, and (F) deoxycholic (DCA) and lithocholic acid (LCA) species. **CA species** is the summation of cholic, glycocholic and taurocholic acid; **CDCA species** is the summation of chenodeoxycholic, glycochenodeoxycholic and taurochenodeoxycholic acid; **DCA species** is the summation of deoxycholic, glycodeoxycholic and taurodeoxycholic acid; **LCA species** is the summation of lithocholic and glycolithocholic acid. Variables were log and/or square root transformed prior to repeated measures ANOVA to meet normality and homogeneity of variance assumptions. Total primary bile acids (BA), CA species and CDCA species data points were winsorized at the 95th percentile for data visualization purposes only. Avocado = AV; Control = AA; Nutrients = OF

Total secondary bile acids in the AV group were 34% and 24% lower compared to OF (p < 0.001) and AA (p = 0.011), respectively (**Supplemental Table 4; Figure 2D**). Likewise, total bile acid concentrations in the AV group were 32% and 20% lower compared to OF (p < 0.001) and AA (p = 0.04) (Supplemental Table 4; **Figure 2B**), primarily driven by secondary bile acids concentrations. There were no differences between AA and OF conditions. Lastly, total primary bile acids tended to be higher in both AV and OF groups compared to AA (p = 0.063) (Supplemental Table 4; **Figure 2C**).

Next, we grouped the primary and secondary bile acids along with their glycine and taurine conjugated forms, as previously described (29,30). Within secondary bile acid species, deoxycholic acid (DCA) species concentration was 38% higher in the OF group (p < 0.001) and 24% higher in the AA (p = 0.037) compared to AV (Supplemental Table 4; **Figure 2F**). LCA species concentration was 21% higher in the OF group (p = 0.055) and 19% higher in the AA (p = 0.055) compared to AV (Supplemental Table 4; Figure 2F). Similarly, both DCA (β = -0.12, p = 0.02]) and LCA (β = -0.12, p = 0.03) species were lower in AV compared to AA, while DCA (β = 0.11, p = 0.05) species tended to be higher in OF, compared to AA (Supplemental Table 2). Lastly, there was a trend towards lower UDCA species concentrations in the AV group (p = 0.07) (Supplemental Table 4).

Within primary bile acid species, chenodeoxycholic acid (CDCA) species tended to be higher in both OF and AV groups, compared to AA (p = 0.062). Interestingly, for the CA species, OF had 63% higher concentrations than AA (p = 0.041) (Supplemental Table 4; **Figure 2E**). Despite statistical significance, the OF treatment showed no meaningful association (small effect size) on cholic acid concentrations (β = 0.05 [p = 0.20]) (Supplemental Table 2). Notably, AV also had higher CA species concentration, but this effect was not statistically significant compared to AA. There were no statistically significant differences among the concentrations of HCA species among the groups (p = 0.87) (Supplemental Table 4).

### Gut microbiome composition and function

#### Differential abundance at the genus and species level

There were no differences in the concentrations of *Faecalibacterium* in both AV (β = - 0.04, q = 0.86) and OF (β = 0.07, q = 0.79), compared to AA (**Table 5**; **Figure 3A**). At the genus level, the OF had a higher relative abundance of *Lachnospira* (β = 1.29, q = 0.03), compared to AA. Furthermore, the AV group (β = 0.95, q = 0.05) had a higher relative abundance of *Candidatus_Cibionibacter,* whereas OF (β = -0.82, q = 0.05) had a lower relative abundance of this genus, compared to AA. Notably, there was a numerically higher abundance of *Bacteroides* and *Parabacteroides* in both OF and AV, compared to the control, but this was not statistically significant after FDR correction (q ≥ 0.20) (Table 5; Figure 3A). Likewise, the abundance of *Alistipes* and *Clostridium* was 28% and 44% higher in AV compared to the AA, but did not remain statistically significant after FDR correction (q = 0.25). Lastly, the relative abundance of *Phascolarctobacterium* (β = -0.48, q = 0.11) and *Neglectibacter* (β = -1.24, q = 0.11) was significantly lower in AV, compared to AA (Table 5; Figure 3A).

**Figure 3.**
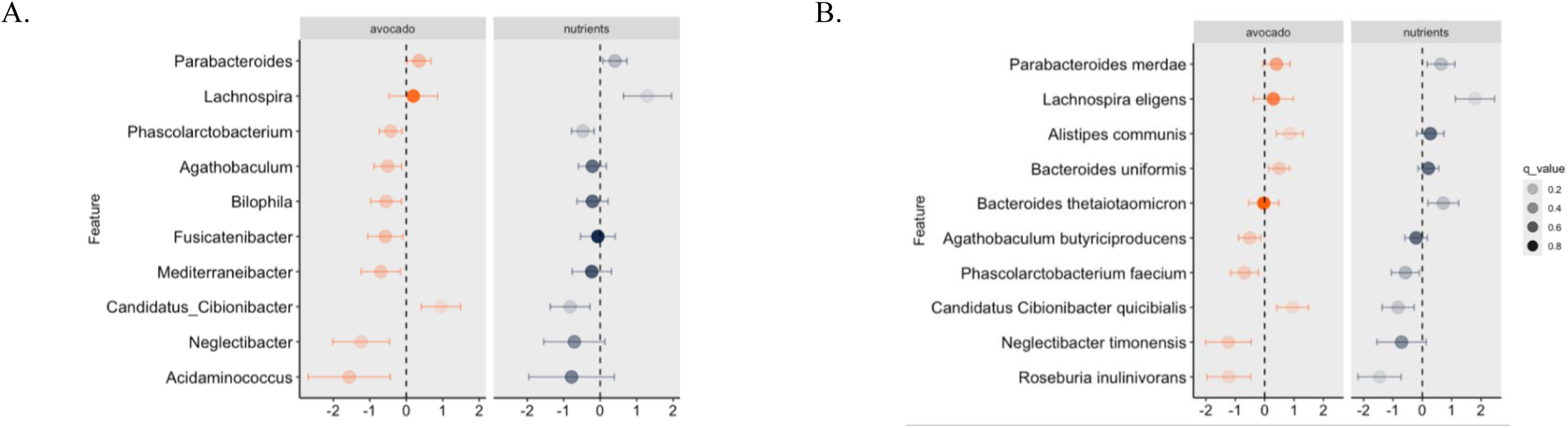
Forest plot of differential abundance analysis at the genus (A) and species (B) level in AV and OF, compared to AA. Differential abundance analysis was performed using MaAsLin3 on the 100 most abundant taxa, retaining those with at least 10 reads per sample and 10% prevalence, resulting in 67 genera and 86 species. Counts were normalized by total sum scaling and scaled to absolute abundances per sample library size. Log-transformed absolute abundances were then analyzed with generalized linear mixed models, including treatment as a fixed effect and participant as a random effect. The x-axis represents the estimated effect size (β coefficient), the vertical dashed line denotes null effect (β = 0), and the horizontal lines denotes the 95% confidence intervals. Significance level for microbial abundance was defined at FDR q ≤0.15. Avocado = AV; Control = AA; Nutrients = OF

**Table 5.** Differential abundance at the genus level across dietary conditions.

| Genus (%) | Control (AA) |  | Nutrients (OF) |  | Avocado (AV) |  | AA-OF |  |  | AA-AV |  |  |
| --- | --- | --- | --- | --- | --- | --- | --- | --- | --- | --- | --- | --- |
|  | Mean | SE | Mean | SE | Mean | SE | Beta (95% CI) | p value | q value | Beta (95% CI) | p value | q value |
| *Faecalibacterium* | 11.3 | 1.08 | 12.4 | 1.22 | 10.9 | 1.03 | 0.07 (-0.21, 0.34) | 0.63 | 0.79 | -0.04 (-0.32, 0.23) | 0.76 | 0.86 |
| *Roseburia* | 3.39 | 0.44 | 3.87 | 0.56 | 2.9 | 0.47 | -0.01 (-0.51, 0.49) | 0.97 | 0.97 | -0.46 (-0.95, 0.03) | 0.07 | 0.31 |
| *Clostridium* | 2.25 | 0.28 | 2.65 | 0.33 | 3.23 | 0.39 | 0.27 (-0.1, 0.65) | 0.15 | 0.45 | 0.42 (0.05, 0.79) | 0.03 | 0.25 |
| *Blautia* | 2.87 | 0.29 | 2.54 | 0.28 | 2.26 | 0.25 | -0.09 (-0.49, 0.29) | 0.62 | 0.79 | -0.36 (-0.75, 0.02) | 0.06 | 0.31 |
| *Ruminococcus* | 2.3 | 0.38 | 1.68 | 0.27 | 2.2 | 0.35 | -0.16 (-0.75, 0.43) | 0.59 | 0.79 | -0.05 (-0.64, 0.54) | 0.86 | 0.89 |
| *Dorea* | 1.45 | 0.21 | 1.28 | 0.17 | 1.07 | 0.13 | -0.11 (-0.53, 0.31) | 0.6 | 0.79 | -0.28 (-0.7, 0.15) | 0.2 | 0.49 |
| ***Lachnospira*** | 0.82 | 0.14 | 2.16 | 0.36 | 1.01 | 0.18 | **1.29 (0.64, 1.96)** | **< 0.001** | **0.03** | 0.19 (-0.47, 0.86) | 0.56 | 0.79 |
| ***Phascolarctobacterium*** | 1.23 | 0.26 | 0.85 | 0.16 | 0.76 | 0.13 | **-0.48 (-0.79, -0.17)** | **< 0.01** | **0.11** | -0.42 (-0.74, -0.12) | < 0.01 | 0.17 |
| ***Candidatus_Cibionibacter*** | 1.04 | 0.25 | 0.77 | 0.24 | 1.5 | 0.25 | **-0.82 (-1.37, -0.28)** | **< 0.01** | **0.11** | **0.95 (0.42, 1.49)** | **< 0.001** | **0.05** |
| ***Neglectibacter*** | 0.25 | 0.09 | 0.22 | 0.08 | 0.14 | 0.04 | -0.71 (-1.55, 0.13) | 0.11 | 0.42 | **-1.24 (-2.01, -0.46)** | **< 0.01** | **0.11** |
| *Mediterraneibacter* | 1.16 | 0.17 | 0.89 | 0.12 | 0.81 | 0.11 | -0.23 (-0.77, 0.31) | 0.4 | 0.69 | -0.7 (-1.24, -0.16) | 0.01 | 0.18 |
| *Fusicatenibacter* | 0.72 | 0.09 | 0.73 | 0.09 | 0.65 | 0.1 | -0.06 (-0.54, 0.41) | 0.79 | 0.88 | -0.57 (-1.05, -0.09) | 0.02 | 0.22 |
| *Acidaminococcus* | 0.34 | 0.17 | 0.24 | 0.11 | 0.24 | 0.13 | -0.79 (-1.96, 0.39) | 0.2 | 0.49 | -1.56 (-2.69, -0.44) | 0.01 | 0.18 |
| *Agathobaculum* | 0.26 | 0.03 | 0.2 | 0.03 | 0.19 | 0.03 | -0.21 (-0.6, 0.17) | 0.28 | 0.57 | -0.51 (-0.88, -0.13) | 0.01 | 0.18 |
| *Bacteroides* | 8.94 | 1.04 | 10.1 | 1.27 | 11.3 | 1.47 | 0.26 (0, 0.52) | 0.05 | 0.26 | 0.26 (0, 0.52) | 0.05 | 0.26 |
| *Alistipes* | 4.25 | 0.52 | 4.59 | 0.54 | 5.42 | 0.62 | 0.21 (-0.07, 0.49) | 0.14 | 0.45 | 0.32 (0.04, 0.6) | 0.02 | 0.25 |
| *Parabacteroides* | 1.79 | 0.24 | 2.19 | 0.29 | 2.12 | 0.27 | 0.4 (0.08, 0.73) | 0.01 | 0.2 | 0.35 (0.02, 0.68) | 0.03 | 0.26 |
| *Prevotella* | 8.09 | 2.03 | 7.23 | 1.8 | 7.15 | 1.98 | 0.16 (-0.61, 0.94) | 0.68 | 0.81 | -0.24 (-1.02, 0.53) | 0.54 | 0.79 |
| *Bifidobacterium* | 1.31 | 0.24 | 1.03 | 0.2 | 1.19 | 0.19 | -0.55 (-1.36, 0.27) | 0.19 | 0.49 | -0.57 (-1.37, 0.22) | 0.16 | 0.45 |
| *Collinsella* | 4.16 | 0.79 | 2.89 | 0.51 | 2.75 | 0.52 | -0.26 (-0.93, 0.41) | 0.44 | 0.74 | -0.48 (-1.16, 0.19) | 0.16 | 0.45 |
| *Akkermansia* | 1.33 | 0.54 | 1.43 | 0.61 | 1.69 | 0.68 | -0.2 (-0.94, 0.55) | 0.6 | 0.79 | 0.33 (-0.41, 1.08) | 0.38 | 0.67 |
| *Bilophila* | 0.29 | 0.04 | 0.22 | 0.04 | 0.18 | 0.03 | -0.21 (-1.95, 0.21) | 0.33 | 0.62 | -0.55 (-2.30, -0.13) | 0.01 | 0.18 |

Values are means ± standard error. For differential abundance of metagenomics data, analysis was performed with the Microbiome Multivariable Associations with Linear Models 3 (MaAsLin3) package in R, using a generalized mixed effects model with treatment as a fixed effect and participant ID as a random effect. $\beta$ coefficients are reported with 95% confidence interval in parenthesis (95% CI), raw p values and FDR-adjusted p values (q value).

There were no differences in the differential abundance of *F. prausnitizi* in both AV (β = - 0.06, q = 0.88) and OF (β = 0.03, q = 0.92), compared to AA (**Table 6**; **Figure 3B**). At the species level, the relative abundance of *Lachnospira eligens* (β = 1.79, q <0.001) was significantly higher in OF, compared to AA. Similar to the *Candidatus_Cibionibacter* genus, the OF group (β = -0.82, q = 0.11) had a lower relative abundance of *Candidatus_Cibionibacter quicibialis* species, while AV (β = 0.95, q = 0.03) had a higher relative abundance, relative to the AA. Furthermore, there was a lower relative abundance of *Phascolarctobacterium faecium* (β = -0.69, q = 0.14) and *Neglectibacter timonensis* (β = -1.24, q = 0.14) as well as a higher relative abundance of *Alistipes communis* (β = 0.85, q = 0.03) and *Bacteroides uniformis* (β=0.50, q = 0.14) only in AV, compared to AA (Table 6; Figure 3B). Notably, other species within the *Alistipes* genus, such as *A. putredinis* and *A. shahii*, were 9% and 45% higher in AV, compared to AA, but the results were no longer significant after FDR correction (q ≥ 0.63). Relative to AA, the abundance of *Roseburia inulinivorans* was 42% and 25% lower in both the OF group (q < 0.001) and the AV group (q = 0.07), respectively (Table 6; Figure 3B).

**Table 6.** Differential abundance at the species level across dietary conditions.

| Species (%) | Control (AA) |  | Nutrients (OF) |  | Avocado (AV) |  | AA-OF |  |  | AA-AV |  |  |
| --- | --- | --- | --- | --- | --- | --- | --- | --- | --- | --- | --- | --- |
|  | Mean | SE | Mean | SE | Mean | SE | Beta (95% CI) | P value | q value | Beta (95% CI) | P value | q value |
| *Faecalibacterium prausnitzii* | 10.3 | 0.98 | 11.4 | 1.17 | 10.2 | 1.01 | 0.03 (-0.26, 0.32) | 0.84 | 0.92 | -0.06 (-0.34, 0.23) | 0.70 | 0.88 |
| ***Roseburia inulinivorans*** | 0.55 | 0.11 | 0.41 | 0.13 | 0.32 | 0.09 | **-1.46 (-2.19, -0.72)** | **< 0.001** | **0.02** | **-1.21 (-1.96, -0.47)** | **< 0.001** | **0.07** |
| *Roseburia intestinalis* | 0.39 | 0.11 | 0.66 | 0.19 | 0.35 | 0.11 | -0.08 (-0.88, 0.71) | 0.83 | 0.92 | 0.07 (-0.73, 0.88) | 0.85 | 0.92 |
| *Clostridium fessum* | 0.34 | 0.06 | 0.35 | 0.05 | 0.26 | 0.04 | 0.36 (-0.05, 0.77) | 0.09 | 0.41 | -0.02 (-0.42, 0.39) | 0.92 | 0.95 |
| *Clostridium sp AM49 4BH* | 0.29 | 0.07 | 0.38 | 0.11 | 0.52 | 0.14 | 0.06 (-0.82, 0.95) | 0.88 | 0.94 | 0.64 (-0.24, 1.52) | 0.16 | 0.51 |
| *Blautia obeum* | 0.5 | 0.10 | 0.41 | 0.09 | 0.45 | 0.10 | -0.14 (-0.65, 0.37) | 0.58 | 0.83 | -0.10 (-0.63, 0.42) | 0.70 | 0.88 |
| *Ruminococcus torques* | 0.78 | 0.13 | 0.65 | 0.12 | 0.50 | 0.08 | -0.21 (-0.76, 0.33) | 0.44 | 0.74 | -0.69 (-1.25, -0.14) | 0.01 | 0.19 |
| ***Lachnospira eligens*** | 0.32 | 0.06 | 1.00 | 0.18 | 0.33 | 0.07 | **1.79 (1.13, 2.46)** | **<0.001** | **<0.001** | 0.26 (-0.38, 0.97) | 0.39 | 0.70 |
| *Lachnospira pectinoschiza* | 0.41 | 0.11 | 1.02 | 0.30 | 0.60 | 0.17 | 0.84 (0.21, 1.46) | 0.01 | 0.17 | 0.002 (-0.62, 0.63) | 0.99 | 0.99 |
| ***Phascolarctobacterium faecium*** | 0.91 | 0.21 | 0.63 | 0.15 | 0.53 | 0.12 | -0.58 (-1.05, -0.10) | 0.02 | 0.21 | **-0.69 (-1.17, -0.21)** | **<0.05** | **0.14** |
| ***Candidatus Cibionibacter quicibialis*** | 1.04 | 0.25 | 0.77 | 0.24 | 1.50 | 0.25 | **-0.82 (-1.37, -0.28)** | **<0.01** | **0.11** | **0.95 (0.42, 1.49)** | **<0.001** | **0.03** |
| ***Neglectibacter timonensis*** | 0.25 | 0.09 | 0.22 | 0.08 | 0.14 | 0.04 | -0.71 (-1.55, 0.13) | 0.11 | 0.44 | **-1.24 (-2.01, -0.46)** | **<0.01** | **0.11** |
| *Eubacterium rectale* | 4.90 | 0.81 | 4.53 | 0.89 | 2.89 | 0.52 | -0.003 (-0.63, 0.62) | 0.99 | 0.99 | -0.62 (-1.24, -0.02) | 0.04 | 0.29 |
| ***Bacteroides uniformis*** | 3.18 | 0.42 | 3.41 | 0.51 | 4.37 | 0.59 | 0.21 (-0.15, 0.56) | 0.25 | 0.63 | **0.50 (0.14, 0.85)** | **<0.05** | **0.14** |
| *Bacteroides intestinalis* | 0.06 | 0.04 | 0.14 | 0.06 | 0.22 | 0.12 | 0.60 (-0.87, 2.07) | 0.45 | 0.74 | 1.43 (-0.07, 2.92) | 0.11 | 0.44 |
| *Bacteroides cellulosilyticus* | 0.25 | 0.11 | 0.27 | 0.10 | 0.20 | 0.09 | 0.14 (-0.55, 0.84) | 0.68 | 0.88 | -0.52 (-1.22, 0.17) | 0.15 | 0.51 |
| *Bacteroides thetaiotaomicron* | 0.31 | 0.09 | 0.4 | 0.10 | 0.34 | 0.12 | 0.71 (0.19, 1.24) | 0.01 | 0.15 | -0.03 (-0.54, 0.48) | 0.91 | 0.95 |
| *Alistipes putredinis* | 1.98 | 0.27 | 2.05 | 0.27 | 2.16 | 0.31 | 0.14 (-0.15, 0.43) | 0.34 | 0.63 | 0.15 (-0.14, 0.44) | 0.31 | 0.63 |
| *Alistipes shahii* | 0.51 | 0.09 | 0.58 | 0.11 | 0.74 | 0.15 | 0.04 (-0.35, 0.43) | 0.83 | 0.92 | 0.32 (-0.06, 0.71) | 0.10 | 0.42 |
| ***Alistipes communis*** | 0.15 | 0.03 | 0.16 | 0.03 | 0.25 | 0.05 | 0.27 (-0.19, 0.74) | 0.25 | 0.63 | **0.85 (0.40, 1.31)** | **<0.001** | **0.03** |
| *Parabacteroides distasonis* | 0.94 | 0.23 | 0.96 | 0.15 | 1.10 | 0.25 | 0.31 (-0.02, 0.64) | 0.07 | 0.37 | 0.30 (-0.03, 0.64) | 0.07 | 0.37 |
| *Parabacteroides merdae* | 0.74 | 0.11 | 1.10 | 0.2 | 0.93 | 0.14 | 0.64 (0.16, 1.11) | 0.01 | 0.15 | 0.40 (-0.07, 0.87) | 0.09 | 0.42 |
| *Prevotella copri clade A* | 5.75 | 1.46 | 5.00 | 1.40 | 5.40 | 1.67 | -0.31 (-1.07, 0.44) | 0.42 | 0.71 | -0.45 (-1.20, 0.30) | 0.24 | 0.63 |
| *Bilophila wadsworthia* | 0.27 | 0.05 | 0.20 | 0.04 | 0.17 | 0.03 | -0.23 (-0.70, 0.24) | 0.33 | 0.63 | -0.55 (-1.02, -0.09) | 0.02 | 0.21 |

Values are means ± standard error. For differential abundance of metagenomics data, analysis was performed with the Microbiome Multivariable Associations with Linear Models 3 (MaAsLin3) package in R, using a generalized mixed effects model with treatment as a fixed effect and participant ID as a random effect. $\beta$ coefficients are reported with 95% confidence interval in parenthesis (95% CI), raw p values and FDR-adjusted p values (q value).

#### Differential abundance of bile acid, SCFA, and carbohydrate (CHO) metabolism genes

##### Bile acid metabolism

There were no statistical differences in bile salt hydrolase (BSH) gene abundance between AV or OF, compared to AA. Yet, species-level stratification of gene abundance showed that *Blautia obeum* contributed less to BSH abundance in AV (β = -1.00, p = 0.03), compared to the AA. However, the result was no longer statistically significant after FDR correction (q = 0.47). Further, *Bacteroides dorei* had a higher contribution of BSH abundance in the OF (β = 0.56, p = 0.02), compared to AA, potentially related to higher concentrations of secondary bile acids in this treatment group. However, the result was not statistically significant after FDR correction (q = 0.47) (**Figure 4A**)

**Figure 4.**
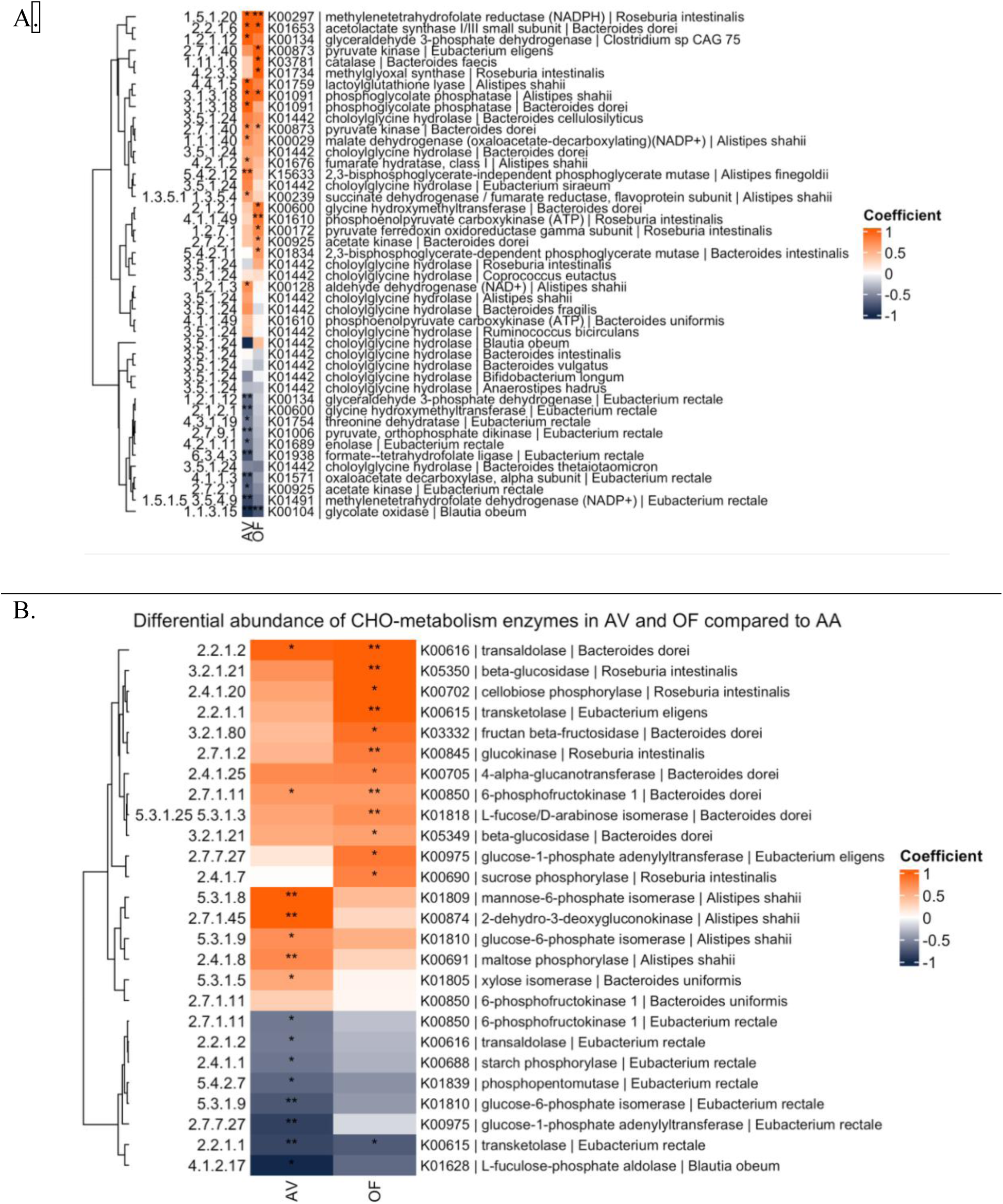
Differential abundance of (A) SCFA and bile acids as well as (B) carbohydrate (CHO) metabolism enzymes. Heatmap displaying gene abundance analysis focusing on pathways involved in (A) short-chain fatty acid and bile acid synthesis (choloylglycine hydrolase), as well as (B) CHO metabolism enzymes using the R package KEGGREST (v1.48.1). Then, to stabilize the generalized linear mixed models for statistical analysis, we filtered for the prevalence and abundance of genes. A total of 16 bile acid–related genes, 1,372 short-chain fatty acid–related 1,138 CHO metabolism enzymes were retained. These filtered data were analyzed using MaAsLin3 to assess differential gene abundance across dietary conditions. Significance threshold q ‘**’ ≤ 0.15, ‘*’ ≤ 0.25. Avocado = AV; Control = AA; Nutrients = OF

##### Short-chain fatty acid (SCFA) metabolism

Acetate is synthesized by gut bacteria via acetyl-CoA synthesis and the Wood-Ljungdahl pathway (31). Through acetyl-CoA formation, pyruvate ferredoxin oxidoreductase catalyzes the oxidative decarboxylation of pyruvate to form acetyl-CoA and CO_2_ (32). Species-level contribution of this enzyme by *Roseburia intestinalis* was higher in both AV and OF, compared to AA; however, it was only statistically significant in the OF group (β = 0.76, q = 0.18) (Figure 4A). Further, this enzyme can indirectly contribute to butyrate synthesis, as butyrate is synthesized by the condensation of two molecules of acetyl-CoA (31). Another important enzyme in acetate synthesis is acetate kinase, which catalyzes the conversion of acetyl-phosphate (derived from acetyl-CoA) to form acetate and ATP (33). Interestingly, the abundance of this enzyme in *Bacteroides dorei* was higher in both AV (β = 0.38) and OF (β = 0.63), but only statistically significant in OF (q = 0.17) (Figure 4A). However, the contribution of this species to pyruvate kinase abundance was statistically higher in both AV (β = 0.74, q = 0.16) and OF (β = 0.60, q = 0.18), which relates to a similar production rate of pyruvate, which is needed to synthesize acetate and the remaining SCFAs (31). On the other hand, the abundance of acetate kinase by *Eubacterium rectale* was lower in both AV (β = -0.71) and OF (β = -0.50), but only statistically significant in AV (q = 0.16) (Figure 4A).

Within the Wood-Ljungdahl pathway, CO_2_ is either reduced to formate or reduced to carbon monoxide, with subsequent reduction with a methyl group to form acetate (31,32). In the “eastern or methyl branch” of the pathway, CO_2_ is reduced to formate, which is subsequently converted to acetyl-CoA through a combination of enzymes such as methylene tetrahydrofolate (THF) reductase and dehydrogenase (34). Interestingly, the species-level contribution of *Roseburia intestinalis* of methylene THF reductase was higher in both AV (β = 1.23, q = 0.18) and OF (β = 1.36, q = 0.14) (Figure 4A). Further, the abundance of methylene THF dehydrogenase in *Eubacterium rectale* was lower in both AV (β = -0.84) and OF (β = -0.58), but only statistically significant in AV (q = 0.14). Propionate is mainly synthesized via the succinate pathway through the conversion of succinate to methyl malonyl-CoA, as well as via the acrylate pathway (using lactate) and the propanediol pathway (using fucose and rhamnose sugars) (31). In the AV group, the species-level contribution of succinate pathway genes by *Alistipes shahii,* such as succinate dehydrogenase (β = 0.61, q = 0.21), fumarate hydratase (β = 0.63, q = 0.18), and malate dehydrogenase (β = 0.71, q = 0.22), was higher compared to AA (Figure 4A). A similar trend was observed in OF, but the results were not statistically significant. Altogether, the results for differential gene abundance in SCFA metabolism suggest that AV and OF have similarities in functional potential, with differences in the species-level contribution of such genes (Figure 4A).

##### Carbohydrate metabolism

Analysis of carbohydrate metabolism gene abundance stratified by species revealed that *Alistipes shahii* within the AV group harbored high levels of enzymes involved in glycolysis such as glucose-6-phosphate isomerase (β = 0.72, q = 0.20) and mannose-6-phosphate isomerase (β = 0.99, q = 0.10) as well as starch and sucrose metabolism enzymes like maltose phosphorylase (β = 0.76, q = 0.10), compared to AA (**Figure 4B**). Interestingly, the abundance of enzymes related to pectin degradation, such as 2-dehydro-3-deoxygluconate (KDG) kinase (β = 1.10, q = 0.10), was also higher in *Alistipes shahii* within AV, compared to AA. Pectin can be degraded by gut microbes, producing oligosaccharides of galacturonic and glucuronic acids, which are then converted to the key intermediate KDG (35,36). KDG is phosphorylated by KDG kinase, and the product of this reaction is cleaved by an aldolase, producing pyruvate and glyceraldehyde-3-phosphate via the Entner–Doudoroff pathway. This pathway can be utilized by bacteria for energy production (35,36). Furthermore, the differentially abundant *Bacteroides uniformis* in the AV group had a higher abundance of xylose isomerase (β = 0.56, q = 0.22) compared to the AA (Figure 4B). The degradation of xylans and xyloglucans, which are hemicelluloses found in avocados (15), results in xylose molecules that are phosphorylated by xylose isomerase to synthesize xylulose (37). Through a series of other enzymatic steps, xylulose can enter the pentose phosphate pathway, potentially serving as a building block for anabolic pathways in *Bacteroides uniformis*.

On the other hand, the OF group showed a high abundance of enzymes involved in cellulose metabolism (38,39), such as cellobiose phosphorylase (β = 1.06, q = 0.15) and β-glucosidase (β = 1.06, q = 0.10), stratified by *Roseburia intestinalis* (Figure 4B) In the AV group, a similar trend was observed, but the results were not statistically significant. Within the same species, sucrose phosphorylase (β = 0.80, q = 0.21) and glucokinase were higher (β = 0.82, q = 0.10) in OF compared to AA, which could relate to energy production through cellulose degradation. *Bacteroides dorei* in the OF group also had a higher abundance of β-glucosidase (β = 0.61, q = 0.20), as well as enzymes such as fructan β-fructosidase, which hydrolyzes the terminal bond of inulin to release fructose (40). Furthermore, these species had a higher abundance of L-fucose/D-arabinose isomerase that catalyzes the conversion of L-fucose to L-fuculose and D-arabinose to D-ribulose, respectively (41).

The major source of L-fucose in adults is fucosylated glycans produced by intestinal epithelial cells (42). The L-fucose can be hydrolyzed and used by gut microbes such as *Bacteroides* genera, to synthesize SCFAs, particularly acetate (42), through the fucose-1-phosphate pathway. The enzyme L-fucose isomerase, involved in the first step of the pathway, was higher in OF (β = 0.72, q = 0.14), compared to AA. However, the enzyme L-fuculase phosphate aldolase stratified by *Blautia obeum*, which converts L-fuculose-1-phosphate to dihydroxyacetophenone (DHAP) (intermediate in the glycolysis pathway) in the third step of the pathway, significantly lower in AV (β = -1.05, q = 0.22), compared to AA. The main source of D-arabinose is found in the side chains of pectins such as rhamnogalacturonan-1 (RG-1) (43,44). The abundance of D-arabinose isomerase was higher in OF (β = 0.72, q = 0.14), compared to AA. The results suggest that the microbiome of the OF is more likely to utilize both external (dietary fiber) and internal (host) carbohydrate sources for energy production and acetate synthesis, whereas the AV is more likely to utilize dietary fiber. Lastly, similar to SCFA metabolism enzymes, enzymes stratified by *Eubacterium rectale,* such as 6-phosphofructokinase (β = -0.59, q = 0.22), transaldolase (β = - 0.58, q = 0.22), starch phosphorylase (β = -0.60, q = 0.21), phosphopentomutase (β = -0.67, q = 0.15), glucose-6-phosphate isomerase (β = -0.75, q = 0.05) and glucose-1-phosphate adenvlyltransferase (β = -0.86, q = 0.10), in the AV were significantly lower, compared to AA (Figure 4B).

##### Carbohydrate-active enzymes (CAZymes)

Similar to the CHO metabolism enzymes, *Alistipes shahii* exhibited a higher functional potential for degrading dietary fibers, such as mannans and galactomannans in the AV condition. For instance, the enzyme β-mannosidase (β = 0.86, q = 0.09) and α-galactosidase (β = 0.84, q = 0.18) were higher in AV, compared to AA (**Figure 5**). Both enzymes work by hydrolyzing the oligosaccharides of galactomannans, which are produced by the initial enzymatic action of the endo-1,4-β-mannanase enzyme (45). β-mannosidase is known to hydrolyze the β-1,4-linked mannose residues from the non-reducing end of the galactomannan oligosaccharides, while α-galactosidase hydrolyses the terminal α-1,6-linked galactose residues from galactomannans or their oligosaccharides (45,46). Additionally, this species exhibited a higher abundance of the enzyme α-1,2-mannosidase (β = 0.74, q = 0.15), also known as glycoside hydrolase (GH) 92, which can be upregulated in the presence of dietary glycoproteins and plant polysaccharides rich in mannose (46).

**Figure 5.**
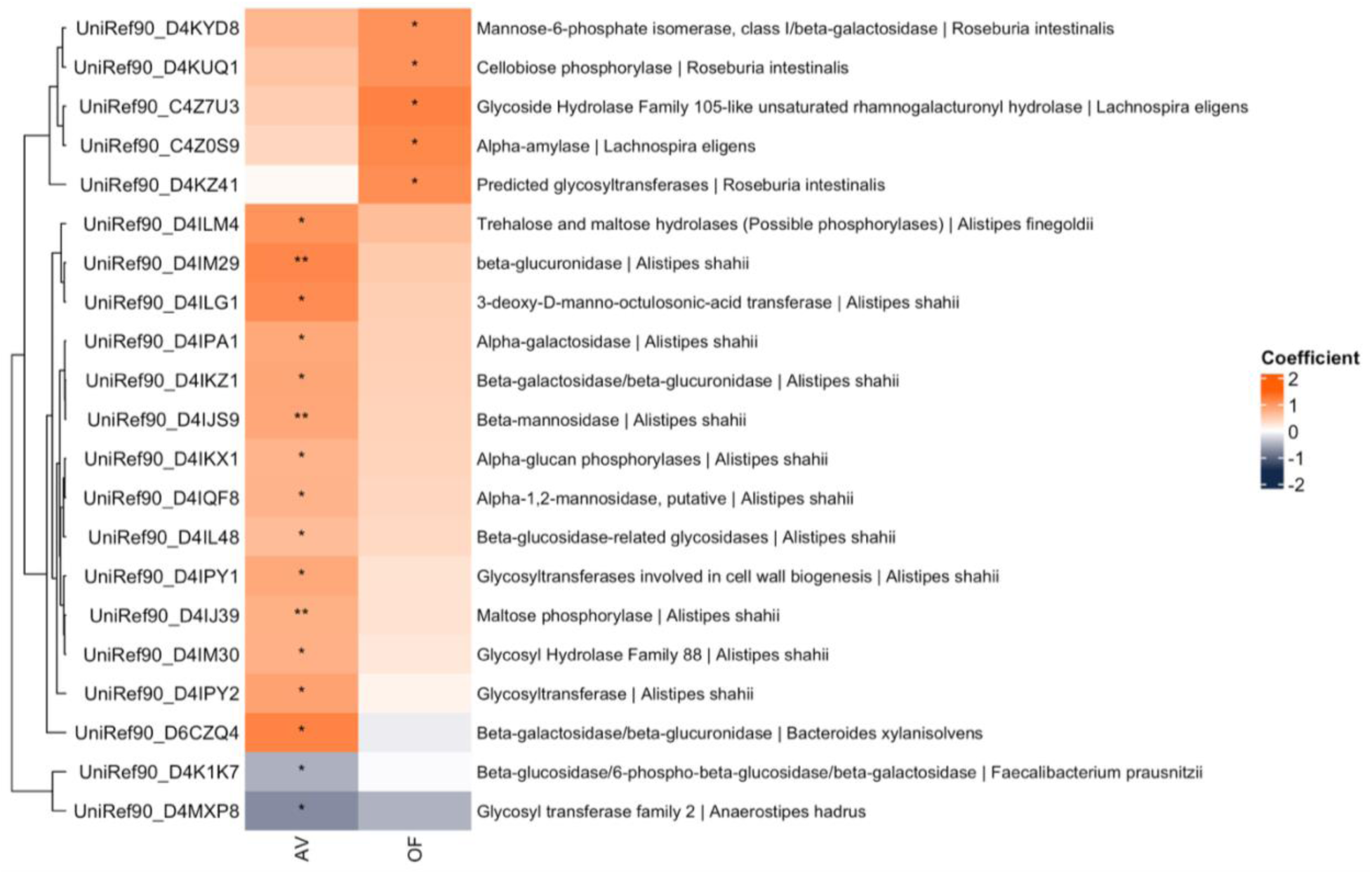
Differential abundance of carbohydrate-active enzymes (CAZymes) in AV and OD, compared to AA. Heatmap displaying gene abundance analysis of CAZymes was conducted was conducted using UniRef 90 annotations stratified by species. To stabilize the generalized linear mixed models for statistical analysis, we filtered using prevalence and abundance thresholds of genes. Of the 1921 CAZymes in our data set, a total of 905 CAZymes met our filtering criteria. These filtered data were analyzed using MaAsLin3 to assess differential gene abundance across dietary conditions. Significance threshold q ‘**’ ≤ 0.15, ‘*’ ≤ 0.25. Avocado = AV; Control = AA; Nutrients = OF

Notably, the abundance of β-glucuronidase (β = 1.15, q = 0.08) was also significantly higher in this species within the AV group (Figure 5). This enzyme is involved in the hydrolysis of exogenous β-glucuronides from the diet or endogenous β-glucuronides produced by the liver, which aids in the detoxification of compounds such as xenobiotics, drugs, and steroids facilitating their adequate excretion in bile through the gastrointestinal tract (47). Gut bacteria can remove the glucuronic acid and affect the enterohepatic circulation of such compounds. Despite this, the enzyme can also play a role in making substrates such as flavonoids (47), more accessible to gut microbes.

Furthermore, *Alistipes shahii* also had a higher abundance of β-glucosidase (β = 0.65, q = 0.24), which is involved in cellulose metabolism, a dietary fiber of which avocados are also a rich source (48,49) as well as enzymes related to starch and lactose metabolism such as α-glucan phosphorylases (β = 0.73, q = 0.17), maltose phosphorylase (β = 0.76, q = 0.09) and β-galactosidase (β = 0.86, q = 0.15) (Figure 5), potentially aiding on metabolizing nutrients that escaped digestion and absorption in the small intestine. Lastly, this species had a higher abundance of enzymes needed for cell wall synthesis and metabolism such as 3-deoxy-D-manno-octulosonic-acid transferase (β = 1.11, q = 0.15) for lipid polysaccharide biosynthesis and glycosyltransferases (GT) family 2 (β = 0.84, q = 0.19) and 4 (β = 0.90, q = 0.18) (Figure 5), suggesting that *Alistipes shahii* thrives in an environment rich of dietary fiber and unsaturated fat such as avocados.

Interestingly, other species within the AV group such as *Bacteroides xylanisolvens,* had higher abundance of β-galactosidase/β-glucuronidase (β = 1.19, q = 0.15), compared to AV, whereas *Faecalibacterium prausnitzii* exhibited a lower abundance of β-galactosidase/β-glucosidase (β = -0.50, q = 0.20). Further, the species *Anaerostipes hadrus* had a lower abundance of GT2 (β = -0.75, q = 0.19), in contrast to *A. shahii* (Figure 5). These results suggest that both *A. shahii* and *B. xylanisolvens* have a higher potential to degrade similar carbohydrates and dietary compounds in the AV group, whereas other species, such as *F. prausnitzii,* do not.

Parallel to carbohydrate metabolism enzymes, the OF group had higher abundance of CAZymes stratified by *Roseburia intestinalis* such as cellobiose phosphorylase (β = 1.05, q = 0.19) and mannose-6-phosphate isomerase (β = 1.06, q = 0.15). Mannose-6-phosphate isomerase, also called phosphomannose isomerase, catalyzes the conversion of mannose-6-phosphate into fructose-6-phosphate during the degradation of the hemicellulose β-mannans (50). Lastly, the abundance of predicted glycosyltransferases (β = 1.08, q = 0.15), involved in cell wall synthesis, was also higher in this species compared to the control (AA) (Figure 5). Furthermore, the differentially abundant *Lachnospira eligens* had a higher abundance of glycoside hydrolase (GH) 105-like rhamnogalacturonyl hydrolase (β = 1.19, q = 0.20) and α-amylase (β = 1.15, q = 0.18). GH 105 acts on RG-1 pectin to release galacturonic acid units (51), thereby enabling pectin degradation, whereas α-amylase catalyzes the hydrolysis of α-1,4-glucose bonds in starch and related α-glucans (52). Altogether, these results suggest *Roseburia intestinalis* has the potential to degrade both cellulose and mannans, while *Lachnospira eligens* potentially degrades pectin and starch that escapes host digestion and absorption in the nutrients (OF) condition.

### Alpha and beta diversity

Alpha diversity quantifies within sample richness and evennes, whereas beta diversity quantifies between-sample community dissimilarity the diversity (53). Alpha diversity was evaluated using the Shannon index and richness and did not differ among dietary conditions (all p > 0.27) (**Figure 6A**). Median Shannon index were numerically higher in AV group (3.14, IQR: 0.47) compared to AA (3.07, IQR: 0.44) and OF (3.03, IQR: 0.55) (Figure 6A).

**Figure 6.**
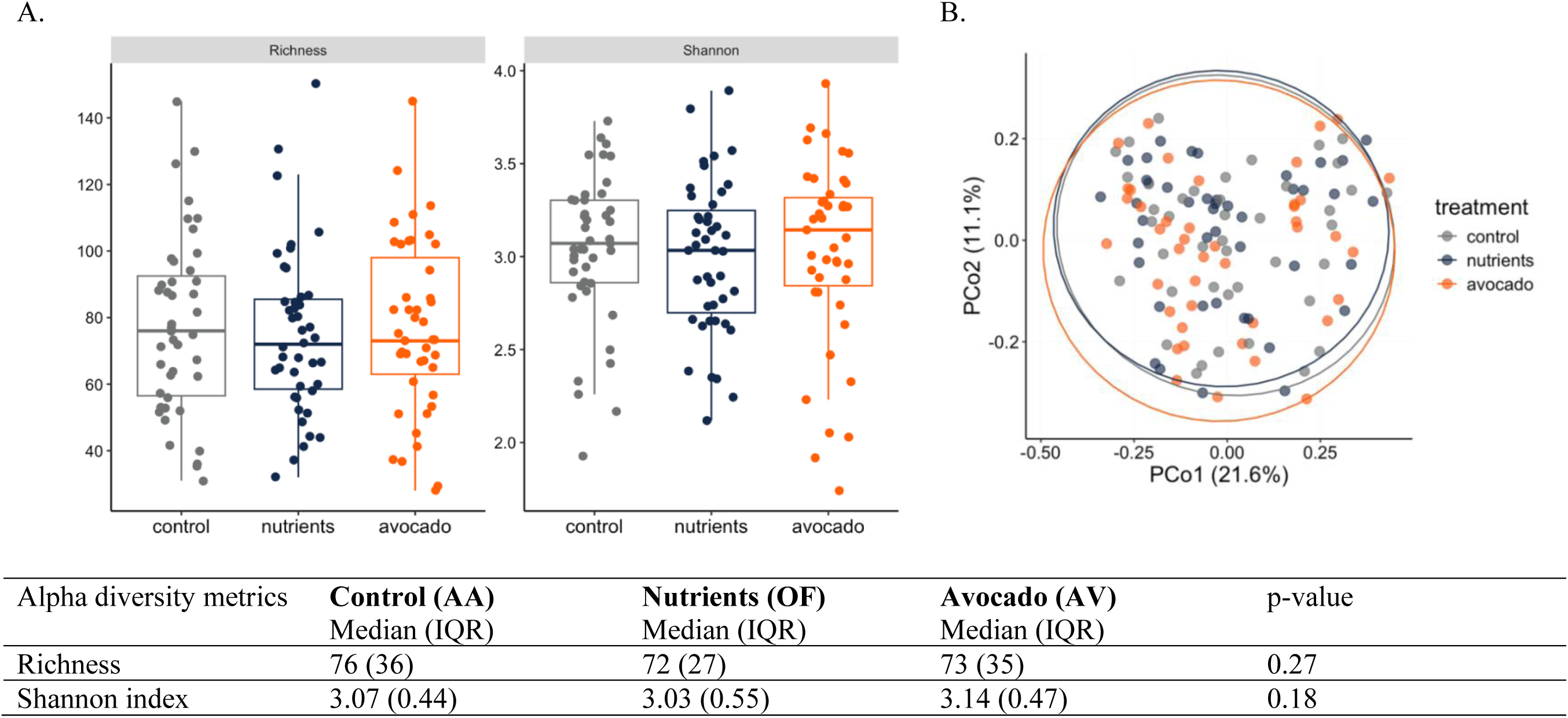
Alpha (A) and Beta diversity (B) measures across dietary conditions. The relative abundance at the genus level was used to calculate alpha (A) and beta (B) diversity. Statistical analysis for Shannon index and Richness was done using non-parametric Friedman test. Principal Coordinate Analysis (PCoA) was done using Bray-Curtis dissimilarity index. Avocado = AV; Control = AA; Nutrients = OF

Community composition, assessed with the Bray-Curtis dissimilarity index, differed across dietary conditions by PERMANOVA (R² = 0.01, p = 0.0014), while homogeneity of dispersion did not differ among the groups (p = 0.93). Pairwise PERMANOVA comparisons were not significant after post hoc testing (all p > 0.65). Consistent with the small effect size (1%), the ordination by Principal Coordinate Analysis (PCoA) (Figure 6B) did not show clear separation by intervention.

### Correlation analysis between omics and microbial species

Correlation analysis utilizing “Hmisc” (v5.2.4) on microbial species and multi-omics data revealed distinct clustering and association patterns with biological relevance. Multi-omics data were clustered in five groups: (1) inflammatory and gut permeability markers, (2) secondary bile acids, (3) total bile acids and diastolic blood pressure, (4) total SCFA and acetate, and (5) primary bile acids **(Figure 7**). Within the first group, the species *L. eligens,* which was significantly enriched in the OF, showed a negative correlation with calprotectin (rho = -0.27, q = 0.01). This result aligns with the lower fecal calprotectin in OF. Further, both *C. Cibionibacter quicibialis* (rho = -0.28, q = 0.01) and *A. shahii* (rho = -0.21, q = 0.01) were negatively associated with sIgA. *C. Cibionibacter quicibialis* was significantly enriched in the AV group, while *A. shahii* exhibited higher functional potential to degrade dietary fibers and produce SCFAs. Altogether, these results suggest lower gut inflammation in the dietary fiber groups, in association with enrichment of specific species (Figure 7).

**Figure 7.**
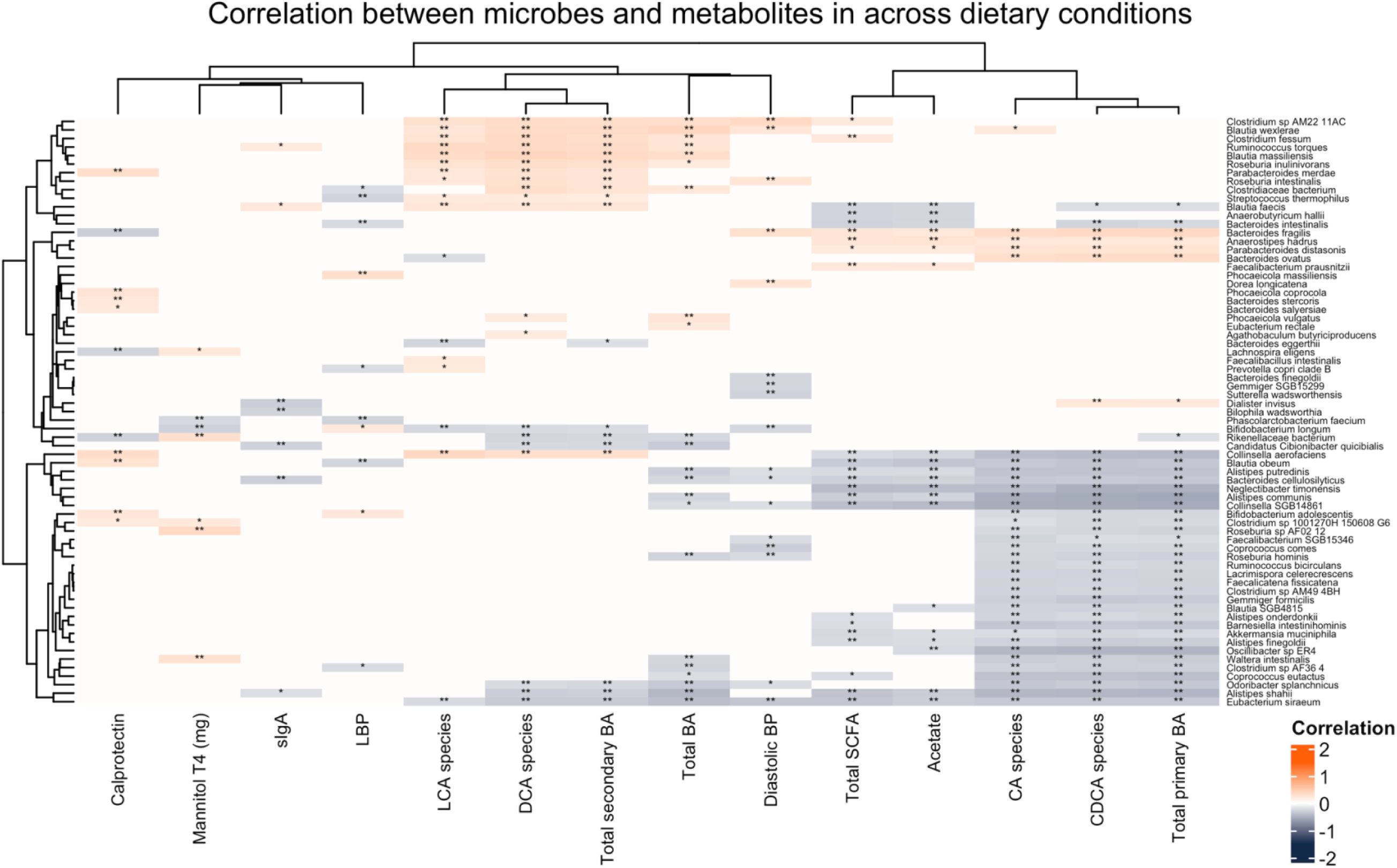
Correlation analysis between microbes and metabolomics data across all dietary conditions. Co-occurrence network analysis was performed using the top 100 most abundant species, along with significant host and microbial phenotypic measures, including fecal and urinary metabolites, inflammatory markers, and anthropometrics from the AV and OF conditions. Spearman correlations were calculated on pooled data across all dietary conditions, with p-values FDR-adjusted (Benjamini–Hochberg, α < 0.10). Networks and clusters were visualized using ComplexHeatmap and circlize. Significance threshold q ‘**’ ≤ 0.05, ‘*’ ≤ 0.1

In the second group, *Clostrium spp.,* such as *C. fessum* and *C. sp AM22 11AC,* were positively correlated with total secondary bile acids, LCA species, and DCA species (all rho ≥ 0.25, q ≤ 0.02). This genus is widely recognized as a key gut microbe related to secondary bile acid synthesis (54), particularly in mechanisms related to deconjugation, as well as oxidation and epimerization of the 3-, 7-, and 12-hydroxy groups of bile acids (55). Furthermore, the species *Ruminococcus torques* was also positively correlated with total secondary bile acids, LCA, and DCA species (all r ≥ 0.34, q ≤ 0.02) (Figure 7). The enzymes 7α/β-HSDHs, involved in epimerization of bile acids, are highly expressed in *Ruminococcus spp* (55). Bile salt hydrolases have also been identified in other genera such as *Bacteroides*, *Blautia*, and *Roseburia* (56).

Interestingly, several *Blautia spp*., including *B. wexlerae*, *B. faecis*, and *B. massiliensis*, were positively associated with total secondary bile acids, lithocholic acid, and deoxycholic acid species (all rho ≥ 0.24, q ≤ 0.03). Further, *R. intestinalis* and *R. inulinovorans* were also positively associated with these metabolites (all rho ≥ 0.22, q ≤ 0.05), while several species of the genus Alistipes, including *A. shahii*, were negatively correlated with these metabolites (all rho ≤ -0.31, q ≤ 0.001) (Figure 7). Although not significantly higher in the differentially abundant analysis, *R. intestinalis* in the nutrients group and *A. shahii* in AV exhibited higher functional potential for SCFA and CHO metabolism enzymes. The observed association between *R. intestinalis* and secondary bile acids may be influenced by the bioaccessibility of both dietary fiber and fat, as higher concentrations of secondary bile acids were detected in OF compared to AV. Conversely, the lower fat bioavailability in avocados could limit secondary bile acid production, potentially explaining the negative correlation observed between *Alistipes* spp. and secondary bile acids.

The third clustering group reflected total BAs and diastolic blood pressure. *A. putredinis* showed a negative correlation with diastolic blood pressure (rho = -0.21, q = 0.09). Furthermore, *R. hominis*, which was reported as a biomarker for normal tension in a cross-sectional study (57), was also negatively correlated with diastolic blood pressure (rho = -0.23, q = 0.04) (Figure 7). Interestingly, species such as *R. intestinalis, B. wexlerea,* and *C. sp AM22 11AC*, that were positively associated with secondary bile acids, were also positively associated with total bile acids and diastolic blood pressure (all rho ≥ 0.25, q = 0.03). Overall, secondary bile acids are recognized as detrimental, as they can increase gut inflammation and intestinal permeability (10,55). However, regarding blood pressure, some studies suggest that secondary bile acids may inhibit or inactivate the reabsorption of sodium in the kidneys, thereby influencing blood pressure (58). Further research is needed to establish a mechanistic or causal relationship between blood pressure and bile acids in feces and circulation on humans.

In the total SCFA and acetate clustering group, there was a positive correlation between total SCFA as well as acetate with *F. prausnitzii, A. hadrus, P. distasonis,* and *B. fragilis* (all rho ≥ 0.21, q ≤ 0.08). Interestingly, *C. fessum* and *C. sp AM22 11AC* were also positively associated with total SCFA (all rho ≥ 0.21, q ≤ 0.07), which could suggest dual ability to synthesize SCFA and secondary bile acids (Figure 7). *F. prausnitzii* is a known gut commensal bacterium, with numerous anti-inflammatory properties in the gut (59). Its health benefits are attributed mainly to its capacity to synthesize butyrate, a major source of energy for colonocytes. Furthermore, *A. hadrus* is also considered a butyrate producer (60), consequently influencing gut physiology. Both microbes engage in cross-feeding, which is the exchange of metabolites (e.g., SCFAs) as energy and nutrients among commensal bacteria (61). For instance, butyrate can be produced by acetate through the acetyl-CoA pathway, as previously described (31,61). These microbes can produce these metabolites as primary degraders in the gut, mainly *Bacteroides, Parabacteroides,* and *Prevotella spp.* contain an array of carbohydrate-active enzymes that help them degrade complex dietary fibers. This can also explain why *P. distasonis* and *B. fragilis* were also positively correlated with acetate, aside from being acetate producers themselves (62,63). Further, other species typically associated with detrimental effects, such as *C. aerofaciens,* and the microbe *N. timonensis*, which was lower in AV compared to AA, showed negative correlations with acetate (all rho ≤ - 0.26, q ≤ 0.03), suggesting a protective role of acetate in maintaining gut microbiota synbiosis. Notably, there was a negative correlation of many *Alistipes spp.,* with acetate and total SCFA (all rho ≤ - 0.22, q ≤ 0.05) (Figure 7). Given the high functional potential of *A. shahii* to produce acetate in AV, it is possible that the species may be synthesizing the metabolite for use by other species through cross-feeding rather than for its own metabolism.

Lastly, the primary bile acids cluster contained several species mainly from the *Firmicutes* phylum, such as *Clostridium, Blautia, Coprococcus, Roseburia, Ruminococcus* and *Anaerostipes*. All these species correlated negatively with total primary, CA, and CDCA species (all rho ≤-0.21, q ≤ 0.08) (Figure 7). Bile acids have the potential to act as antimicrobial agents by damaging and altering the lipids of the cell membrane (64). Interestingly, these species are also gram-positive, and such bacteria generally have the most diverse distribution of bile salt hydrolases (BSH) (64). Furthermore, a study evaluating the role of CA supplementation in regulating the gut microbiota in mice showed a significant increase of Firmicutes spp. over Bacteroidetes with CA consumption in mice (65). Altogether, this exploratory analysis suggests that these microbes possess the ability to resist the antimicrobial potential of bile acids and thrive in the gut environment.

### Systemic and gastrointestinal inflammatory markers

For the present analysis, 41 participants were assessed for AV and OF, and 42 participants for AA. The difference in sample size was due to the inability to collect blood samples. There were no statistically significant differences in any of the systemic inflammatory markers, including LBP (all p > 0.50) (**Table 7**). However, statistically significant differences were observed in the gastrointestinal inflammatory markers measured. Both OF and AV had 26.4% (p = 0.04) and 30.3% (p = 0.03) lower levels of fecal calprotectin, respectively, compared to AA (Table 7; **Supplemental Figure 2A**). Interestingly, when we assessed the effect and magnitude of the relationship between calprotectin and treatment, only AV exhibited a significant negative association (β = -0.11, p = 0.04) (Supplemental Table 2). Furthermore, only the AV group had 34.2% lower levels of secretory immunoglobulin A (sIgA) compared to the control (AA) (p = 0.01), whereas no statistically significant differences were found between OF and AA (p = 0.18), and AV and OF (p = 0.17) (Table 7; **Supplemental Figure 2B**).

**Table 7.** Systemic and gastrointestinal inflammatory markers across dietary conditions.

|  | Control (AA) |  | Nutrients (OF) |  | Avocado (AV) |  | p value |
| --- | --- | --- | --- | --- | --- | --- | --- |
|  | Mean (95% CI) [n] | SEM | Mean (95% CI) [n] | SEM | Mean (95% CI) [n] | SEM |  |
| IL-6, pg/ml<sup>1</sup> | 1.27 (1.07-1.50) [41] | 0.11 | 1.22 (1.03-1.44) [42] | 0.10 | 1.17 (0.98-1.38) [41] | 0.09 | 0.53 |
| TNF-α, pg/ml<sup>2</sup> | 0.83 (0.75-0.91) [41] | 0.04 | 0.81 (0.74-0.90) [42] | 0.04 | 0.83 (0.75-0.91) [41] | 0.04 | 0.89 |
| CRP, mg/L<sup>1</sup> | 2.31 (1.52-3.50) [41] | 0.48 | 2.18 (1.44-3.29) [42] | 0.45 | 2.35 (1.55-3.56) [41] | 0.49 | 0.85 |
| LBP, μg/ml<sup>3</sup> | 12022 (6324.0) [39] | | 11830 (6279.0) [39] | | 12426 (6114.5) [39] | | 0.50 |
| Calprotectin, μg/g<sup>1</sup> | 67.3 (50.3–90.0)<sup>a</sup> [43] | 9.84 | 49.5 (37.0–66.3)<sup>b</sup> [43] | 7.24 | 46.9 (35.0–62.7)<sup>b</sup> [43] | 6.85 | 0.02 |
| sIgA, μg/g<sup>1</sup> | 1655 (1208–2267)<sup>a</sup> [43] | 261 | 1365 (996–1870)<sup>a</sup> [43] | 216 | 1088 (794–1490)<sup>b</sup> [43] | 172 | 0.01 |
| Data are presented as estimated marginal means (EMMs) along with 95% confidence intervals (CI) in parenthesis and sample size [n] in brackets. P-values are derived from linear mixed models (LMMs) with treatment as a fixed effect and participant as the random effect. Variable was log<sup>1</sup> or square root<sup>2</sup> transformed to meet normality and homogeneity of variance assumptions for repeated measures ANOVA. Non-parametric <sup>3</sup>Data are presented as median, interquartile range (IQR) in parenthesis, sample size in brackets [n] and analyzed using the non-parametric Friedman test to variables that did not meet assumptions of normality or homogeneity of variance. Different letters denote differences between groups. Interleukin-6 (IL-6), Tumor necrosis factor alpha (TNF-α), C-reactive protein (CRP), Lipopolysaccharide binding protein (LBP), Secretory Immunoglobulin (sIgA). |  |  |  |  |  |  |  |

### Gut permeability test

Gut permeability was assessed through a non-parametric Friedman test, which requires a complete case design. For timepoint 1 (n = 41), two participants were excluded because they were unable to attend the study visits. Therefore, no data are available across all time points of urine collection. For T2 (n = 38), three additional participants were unable to provide a urine sample within the established time frame. Lastly, in T3 (n = 39), two participants were unable to provide urine samples due to scheduling conflicts. The analysis for the entire time period (time point 4) was done in participants who provided all three previous time points (n = 37) (**Supplemental Tables 5 and 6**). The urinary excretion of orally administered sugar molecules can be used to measure intestinal and colonic permeability in vivo (27). Post-consumption urinary sugar recovery (%) (Supplemental Table 5) and concentrations (mg/mL) (Supplemental Table 6) were analyzed in urine collected at 0-2 hours, 2-8 hours, and 8-24 hours. Overall, there were no statistical differences in the urinary excretion (mg) and recovery (%) of rhamnose, mannitol, and sucralose at timepoints 1, 2, and 3 (Supplemental Tables 5 and 6). Notably, mannitol excretion throughout the entire urine collection (0-24hrs) was 9% and 18% lower in OF and AV, compared AA (p=0.08) (**Figure 8**).

**Figure 8.**
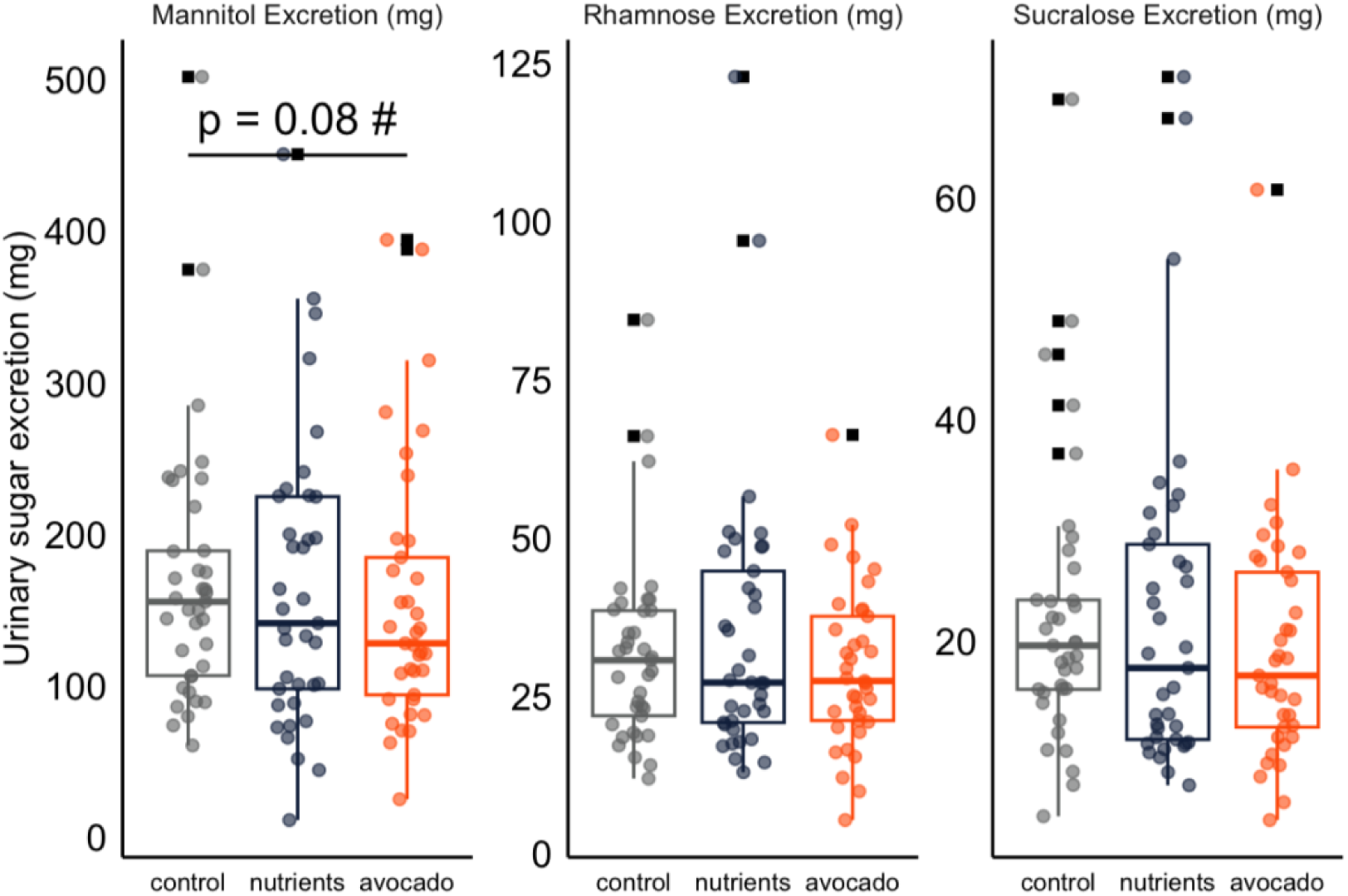
Total (0-24 hrs) urinary sugar cumulative excretion (mg) at each dietary condition. Urinary sugar was quantified through gas chromatography (GC). The data from GC (μg/ml) was used to calculate urinary sugar excretion (mg) by multiplying the sugar (μg/ml) by total urine volume produced (ml) and then dividing by 1000 to obtain milligrams (mg). Statistical analysis was done using non-parametric Friedman test. Avocado = AV; Control = AA; Nutrients = OF

### Gastrointestinal tolerance, regularity, and stool consistency

For consistency purposes, log entries without the required information were classified as missing data, which can influence the sample size for the variables across dietary conditions. The severity of gastrointestinal symptoms reported by participants remained within the absent-to-mild range, with no significant differences observed between dietary conditions for burping, bloating, cramping, nausea, and reflux (all p > 0.16). However, gas severity was higher by 12% and 9% in the AV group compared to AA (p = 0.01) and OF (p = 0.04), respectively, and no differences were observed between the AA and OF conditions (p = 0.48) (**Table 8**). There was a positive trend toward higher rumbling scores in both AV and OF groups, compared to AA (p = 0.06). Stool characteristics, such as self-reported consistency, were based on the Bristol Stool Chart (BSC). Ease of passage, stool consistency, and number of bowel movements per day did not differ between dietary conditions (all p > 0.13) (Table 8).

**Table 8.** Gastrointestinal (GI) symptoms and stool characteristics by treatment.

| GI symptoms | Control (AA) |  | Nutrients (OF) |  | Avocado (AV) |  | P value |
| --- | --- | --- | --- | --- | --- | --- | --- |
|  |  | sample size<br>(n) |  | sample size<br>(n) |  | sample<br>size (n) |  |
| **Flatulence<sup>1</sup>** | 1.75 (1.55-1.94)<br>[0.1]a | 42 | 1.80 (1.60-2.00)<br>[0.1]a | 42 | 1.98 (1.78-2.18)<br>[0.1]b | 41 | < 0.05 |
| Rumbling<sup>2</sup> | 1 (0.24) | 39 | 1 (0.36) | 39 | 1 (0.36) | 39 | 0.06 |
| Burping<sup>2</sup> | 1 (0.39) | 38 | 1 (0.29) | 38 | 1 (0.79) | 38 | 0.76 |
| Bloating<sup>2</sup> | 1 (0.29) | 39 | 1 (0.5) | 39 | 1.14 (0.5) | 39 | 0.16 |
| Cramping<sup>2</sup> | 1 (0.18) | 39 | 1 (0.15) | 39 | 1 (0.21) | 39 | 0.95 |
| Nausea<sup>2</sup> | 1 (0) | 38 | 1 (0) | 38 | 1 (0) | 38 | 0.72 |
| Reflux<sup>2</sup> | 1 (0) | 38 | 1 (0) | 38 | 1 (0) | 38 | 0.24 |
| Bristol Stool Chart<br>(BSC)<sup>1</sup> | 3.65 (3.34-3.96)<br>[0.16] | 42 | 3.83 (3.52-4.14)<br>[0.16] | 42 | 3.63 (3.32-3.95)<br>[0.16] | 41 | 0.19 |
| Stool Ease<sup>1</sup> | 2.32 (2.10-2.53)<br>[0.11] | 41 | 2.18 (1.97-2.40)<br>[0.11] | 41 | 2.35 (2.13-2.56)<br>[0.11] | 40 | 0.13 |
| Bowel Movements<sup>1</sup> | 1.42 (1.19-1.64)<br>[0.11] | 42 | 1.40 (1.18-1.62)<br>[0.11] | 42 | 1.43 (1.20-1.65)<br>[0.11] | 41 | 0.93 |
| pH<sup>1</sup> | 6.97 (6.82-7.12)<br>[0.07] | 42 | 6.82 (6.67-6.96)<br>[0.07] | 43 | 6.89 (6.74-7.03)<br>[0.07] | 43 | 0.11 |

## DISCUSSION

In this clinical trial, daily consumption of avocados as part of a typical American diet supported gastrointestinal health in adults with overweight and obesity. Specifically, daily avocado consumption for 4 weeks, as part of a complete feeding trial, resulted in with higher fecal acetate and total SCFA concentrations, as well as lower fecal calprotectin, sIgA, and secondary bile acid species concentrations. Avocado consumption also enriched *B. uniformis, A. communis,* and *C. cibionibacter quicibalis,* which corresponded to a higher functional potential for fiber metabolism and SCFA stratified by *Alistipes spp*. In contrast, the OF condition exhibited similar levels of acetate and fecal calprotectin to those of avocado but higher fecal secondary bile acids species. Furthermore, microbial enrichment differed, with a higher abundance of *L. eligens* and a functional potential for fiber utilization, stratified by *R. intestinalis, L. eligens* and *B. dorei*.

Previous work from our group reported higher fecal acetate concentrations following 12 weeks of daily avocado consumption (18). In contrast to our previous findings (18) and those of other researchers (66), we did not find an enrichment of *Faecalibacterium spp.* or higher α-diversity in the AV or OF conditions compared to the AA. Notably, Yang et al (66) reported an increase in *F. prausnitzii* abundance and α-diversity in a subset of participants following daily avocado intake (1 large avocado) who also improved their Healthy Eating Index (HEI)-2015 scores from baseline. This suggests that improvements in diet quality, due to increased intake of plant foods, or longer dietary interventions (12-26 weeks) (18,66), might be necessary to promote enrichment of pectin degrader *F. prausnitzii*, as previously described (67,68).

Conversely, *Bacteroides uniformis,* a species with high health-associated core keystone (HACK) index scores, which reflects taxa associated with prevalence in non-diseased individuals, longitudinal stability, and favorable health profiles, was enriched with avocado consumption (69). Species with high HACK index scores are also positively associated with nutrient preferences for xylans and xyloglucan fibers (69). *Bacteroides spp.* are considered primary fiber degraders (61) due to their extensive carbohydrate-active enzyme (CAZyme) repertoire (44). *B. uniformis* can hydrolyze undigested polyphenols in the colon (70) and degrade xyloglucan in vitro (71), of which avocados are a rich source (15). Consistent with this, xylose isomerase attributed to *B. uniformis* was more abundant in AV.

*Alistipes,* a subgenus from the Bacteroidetes (renamed Bacteroidota) phylum, with *A. communis* discovered as recently as 2020 (72), has varying health effects across studies, ranging from associations with animal-based diets (meats, eggs, and cheeses) and increased DCA levels (73) to higher HACK index scores, health benefits, and SCFA production, particularly *A. shahii* and *A. putredinins* (69). In the present study, enhanced propionate production via the succinate pathway by *A. shahii* could have influenced the broader community of taxa with overlapping metabolic niches in the AV condition, such as *P. faecium* (74). Supporting a role in complex CHO utilization, Facimoto et al. reported that *Alistipes spp.* harbor several CAZyme gene clusters (CGCs) in ruminants and humans, that enable degradation of dietary fibers containing mannose, fructose, and glucose sugars (75), in line with higher abundance of α- and β-mannosidase genes stratified by *A. shahii* in the AV condition. Lastly, *Alistipes* abundance has also been reported to increase during dietary patterns rich in polyphenols (68). Collectively, the selective enrichment of *B. uniformis* and *Alistipes* spp. in the AV group may support the utilization of complex, food matrix-embedded fibers in avocados.

In contrast, taxa such as *Roseburia, Eubacterium,* and *Faecalibacterium* often rely on cross-feeding interactions to produce butyrate from acetate, and are generally less specialized for degrading complex carbohydrates (61,76). Consistent with this, the abundance of β-glucosidase and β-galactosidase attributed to *F. prausnitzii*, as well as acetate kinase, 6-phosphofructokinase, and glucose-6-phosphate isomerase by *E. rectale*, was lower in the AV condition, compared to the AA condition. The higher avocado serving size in this study (209g/d), compared with prior trials (∼140-175g/d) (18,66), may have preferentially supported taxa with greater pectin-degrading capacity in the AV condition.

Compared with the AV condition, the OF intervention was associated with higher abundance of CHO- and SCFA-related enzymes stratified to *L. eligens, R. intestinalis,* and *B. dorei*. In line with our results, *L. eligens, R. intestinalis,* and *B. dorei* have been reported to degrade pectic substrates *in vitro*, with *L. eligens* producing a significant concentration of acetate (43). *L. eligens* has been shown to produce acetate in response to other fiber sources, such as psyllium (77), whole grains (78), and almonds (79). These results align with the higher abundance of glycoside hydrolase (GH) 105-like rhamnogalacturonyl hydrolase by *L. eligens* in the OF condition. Interestingly, both AV and OF conditions were associated with lower abundance of *R. inulinivorans* compared to AA, which may reflect this species’ preference for inulin and starch fermentation rather than pectin (80).

Aside from pectin, OF was associated with a higher abundance of enzymes involved in cellulose and hemicelluloses (xylans and mannans) utilization, driven by *R. intestinalis*. In contrast, Chassard et al. reported that *Roseburia* isolates, including *R. intestinalis* cannot degrade pure cellulose, such as microcrystalline cellulose (Avicel ®) (81). However, La Rosa et al reported that *R. intestinalis* harbors polysaccharide utilization loci (PUL) capable of depolymerizing the hemicellulose β-mannan (50). Given that pectin and cellulose were major fiber components in the OF pudding intervention and that almond flour contains hemicelluloses such as xylans and xyloglucan (82), substrates similar to those present in avocados, these fibers may have contributed to the fermentative capacity observed in *R. intestinalis* in OF.

In addition to fiber accessibility, pectin fermentation and acetate production depend on pectin structure (linear vs branched) and degree of methylation (low- vs high-methoxy) (43). The OF pudding contained low-methoxy homogalacturonan pectin, whereas the pectin in ripe avocados is predominantly low-methoxy rhamnogalacturonan-1 pectin (15), which is more branched and potentially less accessible to microbial enzymes. The decision to use homogalacturonan for recipe feasibility of the snacks may have contributed to the distinct species-level stratification of SCFA and CHO metabolism enzymes between fiber groups. Because the food matrix also influences microbial fermentation by enabling or limiting access to key nutrients (14), the structural differences among food items in AV and OF may have contributed to observed differences in bile acid transformation in the colon.

Soluble and viscous fibers, such as pectin, can bind bile acids in the ileum and thereby alter enterohepatic circulation and absorption (2,4). Bile acid synthesis and pool composition are also influenced by dietary fat type (saturated vs. unsaturated) (10) and the food source (dairy fat vs. butter) (83). Unabsorbed bile acids (∼5%), enter the colon where gut microbiota engages in deconjugation (removal of glycine and taurine), hydroxylation (conversion of CA and CDCA to DCA and LCA, respectively), oxidation, and epimerization to form secondary bile acids (8,30,55,64), which are more hydrophobic and detrimental to the cell membranes of the intestinal epithelium (10). As such, secondary bile acids are associated with colon cancer and cholesterol gallstone formation (8). Accordingly, fecal secondary bile acids can serve as a proxy for microbial synthesis in the colon.

Similar to our previous findings (18), avocado intake led to lower concentrations of LCA and DCA species. In contrast, AA and OF had higher levels of LCA species compared to the AV condition, while OF had higher DCA species concentration than both AA and AV. These differences may reflect the physical form of fat in avocados compared with the OF pudding, which can influence CCK release and secretion of primary bile acids (84), and/or the higher bioavailability of primary bile acids in the colon during the OF intervention (2). For instance, CA species were significantly higher in the OF than AA, providing more substrate for DCA synthesis. Consistent with this, Mayengbam et al. reported that pea fiber increases fecal acetate and *L. eligens* abundance, while decreasing CA and DCA (85). Hence, substrate availability can influence the microbial biosynthesis of DCA. Furthermore, *Lachnospira spp.* are primarily involved in the oxidation and epimerization of the 3- (8) and 12-hydroxy (86), rather than the hydroxylation pathways to synthesize DCA from CA. With no significant differences in the abundance of species or enzymes involved in deconjugation (BSH) or hydroxylation (7 α-hydroxylase), it is plausible that higher DCA and LCA in OF are related to higher bio accessibility of fat, rather than increased functional capacity or changes in gut microbiome composition. Altogether, these findings suggest that daily avocado intake may confer protection to the intestinal epithelium by lowering secondary bile acids, despite comparable MUFA and fiber in OF.

Despite higher levels of pro-inflammatory secondary bile acids in the OF condition, both AV and OF conditions lowered fecal calprotectin compared to the AA condition. Fecal calprotectin, a neutrophil-derived inflammatory biomarker in inflammatory bowel disease (IBD) (>100 μg/g), can increase in the setting of gut dysbiosis (87). During intestinal inflammation, neutrophils release calprotectin, which exerts antimicrobial properties by chelating metal ions essential for microbial growth (87). Fiber can alleviate gut inflammation through SCFA by strengthening the mucus layer and supporting immune cell differentiation and maturation (9,10,88). Taxa enriched in AV and OF may have contributed to this anti-inflammatory response. *A. shahii* has been shown to alleviate dextran sulfate sodium (DSS)-induced colitis in a mouse model, with concomitant increases in fecal acetate and propionate (89), and *B. uniformis* abundance improved colitis in a mouse model by increasing mucus layer thickness and decreasing pro-inflammatory cytokines (90). In the OF, the presence of *L. eligens*, an acetate producer with anti-inflammatory properties (43), may have further supported epithelial function.

sIgA, a marker of gut mucosal homeostasis, was lowered in AV compared to the AA condition, and not OF. sIgA is an antibody produced by plasma B cells to prevent colonization of pathogens and translocation of commensal bacteria into the intestinal epithelium (91–93). Furthermore, sIgA binding with probiotics such as *Lactobacillus* and *Bifidobacterium* can enhance their immune function (92). An altered sIgA response, characterized by either excessively high or low concentrations, may be related to diseases such as IBD and metabolic disorders (93). Since gut inflammation was lower, as evidenced by low fecal calprotectin in AV and OF, the body may not have required an active first line of defense in the mucosa, compared to a diet low in MUFA and fiber, such as the AA diet. Future analysis can focus on metagenomic sequencing of bacteria bound to sIgA to determine the role of sIgA, probiotic- or pathogen-focused, in a given environment.

Furthermore, all urinary sugars tested had a lower median excretion (mg/mL) following the 24-hour urine collection in AV and OF, compared to AA, with a trend toward mannitol (p = 0.08). It is reasonable to suggest that the strong gut mucosal barrier (tight junctions) in AV and OF likely limited our ability to detect significant reductions in urinary sugars. A higher dose of the non-caloric sweeteners may have been necessary to detect significant effects, as the dosing in this study was adapted from trials in individuals with gastrointestinal disease, who typically exhibit greater gut permeability (27). Lastly, contrary to our expected outcomes, there was no reduction in circulating lipopolysaccharide binding protein (LBP) in AV, compared to AA and OF. It is possible that systemic inflammatory markers did not shift because the duration of the intervention was insufficient for gut-level improvements to be reflected systemically.

Changes in lesser-known species also distinguished the fiber groups. *C. Cibiobacter quicibialis* was higher in AV but lower in OF, while *N. timonensis* was lower only in AV, compared to AA. Both *N. timonensis* (Taxonomy ID: 1776382) and *C. Cibionibacter quicibalis* (Taxonomy ID: 2500537) belong to the Bacillota phylum, Clostridia class, and Eubacteriales order, with *N. timonensis* classified in the Oscillospiraceae family and formally validated as a genus in 2024 (94). This species has recently been linked to slower gastric transit and mucin-degrading bacteria in individuals with constipation (95). In contrast, *C. Cibiobacter quicibialis* remains an uncultured organism, reflected in its “Candidatus” designation. Notably, a study by Grønbæk et al. found that individuals with higher levels of this species, along with *F. prausnitzii*, clustered together and showed fewer markers of gut inflammation (96). Certainly, further work is needed to enhance taxonomic profiling and understand the roles of these microbes in gut health.

This study has several strengths. The crossover design allowed each participant to serve as their own control, reducing interindividual variability that is common in gut microbiome outcomes (97), while providing adequate statistical power with a smaller sample size and facilitating timely recruitment. The controlled feeding further enhanced internal validity by isolating the biological effects of avocados and their supplemental forms, enabling clear interpretation of food-specific health impact and supporting evidence-based dietary recommendations. Nonetheless, the study also has limitations. Although the 2-week washout aligns with guidelines for clinical trials evaluating gut microbiome (19), a washout period matching the length of the dietary intervention could improve dietary adherence and limit participant fatigue. The controlled feeding design also reduces external validity, and failing to account for participants’ habitual fiber intake may have influenced responsiveness, a limitation that could be overcome by adding a lead-in period (19). Further, natural variation in avocado ripening and post-harvest handling may have introduced fluctuations in nutrient composition across the study period. Lastly, dietary compliance was based on self-reported food logs instead of monitoring intake in a controlled environment, which can lead to recall bias.

In conclusion, adding avocados to a typical American diet was associated with lower gut inflammation, higher acetate and SCFA levels, alongside shifts in microbial taxa and functional potential. Likewise, the MUFA/fiber-matched control pudding (OF) produced similar benefits, though it was accompanied by higher fecal secondary bile acids, which are generally less favorable for colonic health. Thus, consuming whole avocados results in greater benefits to overall health, while offering a cost-effective, sustainable approach to achieve the recommended fiber intake in the U.S. population. The differences between fiber conditions likely reflect variations in the food matrix and nutrient bioavailability to gut microbes. Future studies that account for interindividual differences, including baseline microbiome composition and host determinants of bile acid metabolism, may help clarify mechanisms underlying the differentially observed responses.

## Acknowledgements

NK and HH designed research; MSV, TH, DR, MO, DA and MPT conducted research; MSV, DA, and MPT analyzed data; MSV wrote paper; HH had primary responsibility for final content. All authors have read and approved the final manuscript.

## Data availability

Data described in the manuscript, code book, and analytic code will be made available upon reasonable request, pending approval.

## Declaration of Generative AI and AI-assisted technologies in the writing process

The authors used ChatGPT and Grammarly solely for editorial support to improve clarity, grammar, and readability of author-written text. After using this tool/service, the author(s) reviewed and edited the content as needed and take full responsibility for the content of the publication

## Abbreviations

(MUFA): monounsaturated fatty acids
(AA): Average american
(OF): Oleic acid/fiber nutrients
(AV): avocado
(SCFA): short-chain fatty acids
(BCFA): branched-chain fatty acids
(IL-6): interleukin-6
(CRP): C-reactive protein
(LBP): Lipopolysaccharide binding protein
(TNF-α): Tumor necrosis factor alpha
(DCA species): deoxycholic acid species
(DCA species): lithocholic acid species

