## Supplementary files for "*Persea americana* for Total Health (PATH-2): Effects of Avocado Consumption on Gastrointestinal Health in a Randomized, Crossover, Complete Feeding Trial"

First author: María G. Sanabria-Véaz, R.D.

### Tables

*Supplemental Table 1. Baseline Clinical Characteristics of Participants Randomized to Control (AA), Nutrients (OF), and Avocado (AV)*

| | Overall (n=43) | Control (AA) (n=16) | Nutrients (OF) (n=12) | Avocado (AV) (n=15) |
|---|---|---|---|---|
| Fasting Glucose, mg/dl | 101 ± 6.76 | 101 ± 7.05 | 102 ± 8.20 | 101 ± 5.51 |
| Systolic Blood Pressure, mmHg | 124 ± 14.2 | 121 ± 13.7 | 124 ± 9.06 | 127 ± 17.8 |
| Diastolic Blood Pressure, mmHg | 85 ± 9.84 | 85 ± 11 | 84 ± 7.20 | 86 ± 10.9 |
| ALP, U/L | 64.8 ± 16.6 | 72 ± 19.6 | 63.6 ± 16.7 | 58.2 ± 9.37 |
| ALT, U/L | 28.7 ± 20.9 | 32.3 ± 27.6 | 25.2 ± 13.8 | 27.7 ± 17.7 |
| AST, U/L | 25.8 ± 8.34 | 26 ± 6.4 | 24 ± 5.86 | 27.1 ± 11.5 |

*Baseline Clinical Characteristics of Participants Randomized to Control (AA), Nutrients (OF) and Avocado (AV).* Baseline characteristics of participants randomized and completed the clinical trial (N = 43), stratified by initial treatment assignment. Continuous variables are reported as mean ± standard deviation; categorical variables are reported as number of participants and percentage within each group [n (%)]. Liver enzymes: alkaline phosphatase (ALP), alanine aminotransferase (ALT), aspartate aminotransferase (AST), are provided in units per liter (U/L).

Title: *Persea americana* for Total Health (PATH-2): Effects of Avocado Consumption on Gastrointestinal Health in a Randomized, Crossover, Complete Feeding Trial

First author: María G. Sanabria-Véaz, R.D.

*Supplemental Table 2. Standardized Linear Regression Models Coefficients (β) and 95% Confidence Intervals for Pairwise Differences Between Dietary Conditions*

| | AA-OF | | | AA-AV | | |
|---|---|---|---|---|---|---|
| | β (SE) | 95% CI | p-value | β (SE) | 95% CI | p value |
| Acetate | 0.32 (0.13) | [0.07, 0.58] | *0.01* | 0.51 (0.13) | [0.25, 0.76] | *<0.001* |
| Total Short Chain Fatty Acids | 0.29 (0.13) | [0.04, 0.54] | *0.02* | 0.36 (0.13) | [0.10, 0.61] | *<0.01* |
| Cholic Acid Species | 0.05 (0.04) | [-0.03, 0.13] | 0.20 | -0.01 (0.04) | [-0.09,0.06] | 0.73 |
| Deoxycholic Acid Species | 0.11 (0.05) | [0.00, 0.22] | *0.05* | -0.12 (0.05) | [-0.23, 0.02] | *0.02* |
| Lithocholic Acid Species | 0.01 (0.05) | [-0.09, 0.12] | 0.78 | -0.12 (0.05) | [-0.22, 0.01] | *0.03* |
| Total Primary Bile Acids | 0.06 (0.03) | [-0.01, 0.13] | *0.07* | 0.009 (0.03) | [-0.06, 0.08] | 0.78 |
| Total Secondary Bile Acids | 0.07 (0.05) | [-0.04, 0.17] | 0.20 | -0.12 (0.05) | [-0.22, 0.02] | *0.01* |
| Total Bile Acids | 0.06 (0.04) | [-0.02, 0.15] | 0.13 | -0.08 (0.04) | [-0.16, 0.01] | 0.06 |
| Flatulence | 0.09 (0.12) | [-0.16, 0.34] | 0.48 | 0.36 (0.13) | [0.11, 0.61] | *< 0.01* |
| Diastolic Blood Pressure | -0.08 (0.1) | [-0.28, 0.12] | 0.42 | -0.24 (0.10) | [-0.43, 0.04) | *0.01* |
| Systolic Blood Pressure | 0.05 (0.11) | [-0.17, 0.27] | 0.66 | -0.11 (0.11) | [-0.33, 0.11] | 0.31 |
| Rhamnose 24 hr (mg) | 0.03 (0.06) | [-0.09, 0.15] | 0.63 | -0.02 (0.06) | [-0.14, 0.09] | 0.67 |
| Mannitol 24 hr (mg) | -0.06 (0.07) | [-0.20, 0.08] | 0.36 | -0.05 (0.07) | [-0.19, 0.09] | 0.49 |
| Sucralose 24 hr (mg) | -0.04 (0.06) | [-0.17, 0.09] | 0.57 | -0.06 (0.06) | [-0.19, 0.07] | 0.34 |
| Fecal secretory immunoglobulin A (sIgA) | -0.03 (0.05) | [-0.13, 0.07] | 0.58 | -0.10 (0.05) | [-0.20, -0.01] | *0.03* |
| Fecal calprotectin | -0.09 (0.06) | [-0.20, 0.02] | 0.11 | -0.11 (0.06) | [-0.22, 0.00] | *0.04* |

*Standardized Linear Regression Models Coefficients (β) and 95% Confidence Intervals for Pairwise Differences Between Dietary Conditions* . For variables that were significant ($\alpha \leq 0.05$) or trending toward significance ($\alpha \leq 0.10$), effect sizes were estimated using the model_parameters function from the "parameters" (v0.28.2) package in R. Linear regression models were refit using standardized β with standard error (SE), and 95% confidence intervals are presented in the table and computed using the Satterthwaite method for mixed effects models.

Title: *Persea americana* for Total Health (PATH-2): Effects of Avocado Consumption on Gastrointestinal Health in a Randomized, Crossover, Complete Feeding Trial

First author: María G. Sanabria-Véaz, R.D.

*Supplemental Table 3. Fecal volatile fatty acid, phenols, indoles, and ammonia concentrations by treatment*

| | Control (AA) | | Nutrients (OF) | | Avocado (AV) | | |
|---|---|---|---|---|---|---|---|
| **SCFA (μmol/g)** | Mean (CI) *[n]* | SE | Mean (CI) *[n]* | SE | Mean (CI) *[n]* | SE | p value |
| Acetate | 244 (204-285)[a] *[43]* | 20.3 | 288 (248-328)[b] *[43]* | 20.3 | 312 (272-353)[b] *[43]* | 20.3 | <0.001 |
| Butyrate[1] | 61.0 (50.9-71.9) [43] | 5.27 | 67.1 (56.5-78.6) *[43]* | 5.53 | 60.3 (50.3-71.2) *[43]* | 5.24 | 0.27 |
| Propionate[1] | 72.0 (59.9-85.1) *[43]* | 5.99 | 79.2 (66.5-92.9) *[43]* | 6.77 | 74.7 (62.4-88.0) *[43]* | 6.83 | 0.38 |
| Total | 387 (326-448)[a] [43] | 30.6 | 445 (384-506)[b] *[43]* | 30.6 | 459 (398-520)[b] *[43]* | 30.6 | 0.01 |
| **BCFA (μmol/g)** | | | | | | | |
| Isobutyrate[1] | 7.37 (6.47-8.32) *[43]* | 0.46 | 6.89 (6.02-7.81) *[43]* | 0.45 | 6.97 (6.10-7.89) *[43]* | 0.45 | 0.53 |
| Isovalerate | 10.1 (9.00-11.3) *[43]* | 0.57 | 9.23 (8.10-10.4) *[43]* | 0.57 | 9.6 (8.47-10.7) *[43]* | 0.57 | 0.41 |
| Valerate[2] | 6.82 (5.42-8.57) *[43]* | 0.77 | 7.40 (5.89-9.30) *[43]* | 0.84 | 6.60 (5.25-8.29) *[43]* | 0.75 | 0.23 |
| Total[1] | 25.2 (22.3-28.4) *[43]* | 1.52 | 24.3 (21.4-27.3) *[43]* | 1.49 | 23.8 (21.0-26.9) *[43]* | 1.47 | 0.61 |
| **Phenols (μmol/g)** | | | | | | | |
| 4-methylphenol[1] | 1.91 (1.58-2.27) *[41]* | 0.17 | 1.87 (1.54-2.23) *[40]* | 0.17 | 2.22 (1.85-2.62) *[39]* | 0.19 | 0.15 |
| Total[1] | 2.12 (1.77-2.49) *[42]* | 0.18 | 2.08 (1.74-2.46) *[41]* | 0.18 | 2.21 (1.85-2.59) *[42]* | 0.18 | 0.81 |
| **Indoles (μmol/g)** | | | | | | | |
| Indole[1] | 0.94 (0.79-1.12) *[40]* | 0.08 | 1.09 (0.91-1.29) *[38]* | 0.09 | 0.92 (0.76-1.10) *[39]* | 0.08 | 0.15 |
| Total indole[2] | 1.03 (0.86-1.24) *[42]* | 0.09 | 1.23 (1.02-1.49) *[40]* | 0.11 | 1.26 (1.05-1.53) *[40]* | 0.12 | 0.14 |
| **Ammonia (μmol/g)** | 114 (102-128) *[43]* | 6.66 | 110 (97.6-123) *[43]* | 6.38 | 110 (97.8-123) *[43]* | 6.38 | 0.77 |

*Fecal volatile fatty acid concentrations by treatment.* Estimated marginal means (EMMs) are reported in **μmol/g** dry matter basis (DMB) along with 95% confidence intervals (CI) in parenthesis and sample size [n] in brackets. *P-values* are derived from linear mixed models (LMMs) with treatment as a fixed effect and participant as the random effect. Sqrt[1] or log[2] transformations were done as appropriate to meet normality and homogeneity of variance assumptions for repeated measures ANOVA. Post hoc analysis was performed and adjusted for multiple comparisons using the BH method. Different letters denote differences between groups. SCFA = short chain fatty acids; BCFA = branched chain fatty acids; SEM = standard error of the mean.

Title: *Persea americana* for Total Health (PATH-2): Effects of Avocado Consumption on Gastrointestinal Health in a Randomized, Crossover, Complete Feeding Trial

First author: María G. Sanabria-Véaz, R.D.

*Supplemental Table 4. Bile acids (BA) species concentrations by treatment.*

| | Control (AA) | | Nutrients (OF) | | Avocado (AV) | | |
|---|---|---|---|---|---|---|---|
| **Bile acid (µmol/g)** | Mean (CI) [n] | SEM | Mean (CI) [n] | SEM | Mean (CI) [n] | SEM | p value |
| Total Primary BA[2] | 0.03 (0.01-0.07) *[43]* | 0.01 | 0.07 (0.03-0.17) *[43]* | 0.03 | 0.06 (0.02-0.13) *[43]* | 0.02 | 0.06 |
| Total Secondary BA[2] | 5.2 (4.03-6.71) *[43]*[a] | 0.66 | 5.93 (4.60-7.65) *[43]*[a] | 0.76 | 3.94 (3.05-5.09) *[43]*[b] | 0.5 | < 0.001 |
| Total BA[2] | 6 (4.66-7.74) *[43]*[a] | 0.76 | 7.04 (5.46-9.07) *[43]*[a] | 0.9 | 4.79 (3.71-6.17) *[43]*[b] | 0.61 | < 0.01 |
| **Bile acid species (µmol/g)** | | | | | | | |
| CA species[2] | 0.01 (0.003-0.023)[a] *[43]* | 0.004 | 0.024 (0.009-0.057)[b] *[43]* | 0.01 | 0.014 (0.005-0.033)[a,b] *[43]* | 0.006 | < 0.05 |
| CDCA species[2] | 0.01 (0.005-0.036) *[43]* | 0.006 | 0.033 (0.012-0.089) *[43]* | 0.01 | 0.034 (0.013-0.091) *[43]* | 0.01 | 0.06 |
| HCA species[1] | 0.009 (0.006-0.014) *[43]* | 0.002 | 0.009 (0.006-0.014) *[43]* | 0.002 | 0.01 (0.006-0.015) *[43]* | 0.002 | 0.87 |
| DCA species[1] | 2.90 (2.12-3.80)[a] *[43]* | 0.42 | 3.56 (2.69-4.56)[a] *[43]* | 0.46 | 2.21 (1.54-3.00)[b] *[43]* | 0.37 | < 0.001 |
| DCA species[1] | 2.69 (2.10-3.35)[a] *[43]* | 0.31 | 2.76 (2.17-3.43)[a] *[43]* | 0.31 | 2.17 (1.65-2.77)[b] *[43]* | 0.28 | < 0.05 |
| UDCA species# | 0.011 (0.13) *[43]* | | 0.016 (0.33) *[43]* | | 0.006 (0.08) *[43]* | | 0.07 |

*Bile acid (BA) species concentrations by treatment.* Estimated marginal means (EMMs) are reported in **µmol/g** dry matter basis (DMB) with 95% confidence intervals (CI) in parenthesis and sample size [n] in brackets. *P-values* are derived from linear mixed models (LMMs) with treatment as a fixed effect and participant as the random effect. Sqrt[1] and log[2] transformations were done as appropriate to meet normality and homogeneity of variance assumptions for repeated measures ANOVA. Post hoc comparisons were performed when applicable and adjusted for multiple comparisons using the BH method. #UDCA species is reported as the median and interquartile range (IQR) in parenthesis. Nonparametric Friedman test was performed for UDCA species. The compact letter display (CLD) method was used to denote statistical differences between groups. CA species = cholic acid + glycocholic acid + taurocholic acid; CDCA species = chenodeoxycholic acid + glycochenodeoxycholic acid + taurochenodeoxycholic acid; HCA species = hyocholic acid + hyodeoxycholic acid + glycohyocholic acid + taurohyocholic acid + glycohyodeoxycholic acid; DCA species = deoxycholic acid + glycodeoxycholic acid + taurodeoxycholic acid; LCA species is the summation of lithocholic and glycolithocholic acid; UDCA species = ursodeoxycholic acid + glycoursodeoxycholic acid + tauroursodeoxycholic acid. Different letters denote differences between groups. SEM = standard error of the mean.

Title: *Persea americana* for Total Health (PATH-2): Effects of Avocado Consumption on Gastrointestinal Health in a Randomized, Crossover, Complete Feeding Trial

First author: María G. Sanabria-Véaz, R.D.

*Supplemental Table 5. Urinary sugar recovery (%) and disaccharide to monosaccharide ratio each dietary condition across different timepoints of urine collection*

| **Urinary Sugar (%) Recovery or Ratio** | Control (AA) | | Nutrients (OF) | | Avocado (AV) | | |
|---|---|---|---|---|---|---|---|
| | Median (n) | IQR | Median (n) | IQR | Median (n) | IQR | p value |
| **Timepoint 1 (0-2 hrs)** | | | | | | | |
| Rhamnose | 3.46 (41) | 1.99 | 3.68 (41) | 2.73 | 3.35 (41) | 1.99 | 0.48 |
| Mannitol | 7.24 (41) | 4.75 | 6.49 (41) | 3.93 | 6.91 (41) | 4.81 | 0.46 |
| Sucralose | 0.32 (41) | 0.21 | 0.32 (41) | 0.26 | 0.31 (41) | 0.22 | 0.3 |
| Sucralose/Rhamnose | 0.09 (41) | 0.03 | 0.08 (41) | 0.04 | 0.09 (41) | 0.04 | 0.48 |
| Sucralose/Mannitol | 0.04 (41) | 0.03 | 0.04 (41) | 0.02 | 0.04 (41) | 0.03 | 0.29 |
| **Timepoint 2 (2-8 hrs)** | | | | | | | |
| Rhamnose | 5.29 (38) | 4.8 | 6.34 (38) | 7.71 | 5.97 (38) | 5.24 | 0.22 |
| Mannitol | 10.5 (38) | 6.57 | 11 (38) | 12.3 | 10.2 (38) | 10.6 | 0.35 |
| Sucralose | 0.85 (38) | 0.92 | 0.95 (38) | 1.33 | 0.8 (38) | 0.97 | 0.54 |
| **Timepoint 3 (8-24 hrs)** | | | | | | | |
| Rhamnose | 3.81 (39) | 1.88 | 3.82 (39) | 2.4 | 3.82 (39) | 2.73 | 0.52 |
| Mannitol | 7.61 (39) | 11.4 | 8.72 (39) | 12 | 7.41 (39) | 12.7 | 0.58 |
| Sucralose | 0.49 (39) | 0.37 | 0.45 (39) | 0.26 | 0.46 (39) | 0.38 | 0.54 |
| Sucralose/Rhamnose | 0.13 (39) | 0.09 | 0.13 (39) | 0.06 | 0.13 (39) | 0.08 | 0.78 |
| Sucralose/Mannitol | 0.06 (39) | 0.05 | 0.07 (39) | 0.07 | 0.07 (39) | 0.07 | 0.36 |
| **Timepoint 4 (0-24 hrs)** | | | | | | | |
| Rhamnose | 15.49 (37) | 8.66 | 13.46 (37) | 12.2 | 13.63 (37) | 8.65 | 0.34 |
| Mannitol | 30.7 (37) | 16.8 | 28.3 (37) | 24.8 | 25.9 (37) | 18.5 | 0.13 |
| Sucralose | 1.95 (37) | 0.83 | 1.76 (37) | 1.75 | 1.70 (37) | 1.4 | 0.18 |

*Urinary sugar recovery (%) and disaccharide to monosaccharide ratio each dietary condition across different timepoints of urine collection.* Data are presented as median with sample size (n) in parenthesis and interquartile range (IQR). Statistical analysis was performed using the Friedman nonparametric test.

Title: *Persea americana* for Total Health (PATH-2): Effects of Avocado Consumption on Gastrointestinal Health in a Randomized, Crossover, Complete Feeding Trial

First author: María G. Sanabria-Véaz, R.D.

*Supplemental Table 6. Urinary sugar excretion (mg) and disaccharide to monosaccharide ratio each dietary condition across different timepoints of urine collection*

| **Urinary Sugar (mg) Excretion or Ratio** | Control (AA) | | Nutrients (OF) | | Avocado (AV) | | |
|---|---|---|---|---|---|---|---|
| | Median (n) | IQR | Median (n) | IQR | Median (n) | IQR | p value |
| **Timepoint 1 (0-2 hrs)** | | | | | | | |
| Rhamnose | 6.9 (41) | 4.07 | 7.42 (41) | 5.55 | 6.7 (41) | 4.04 | 0.24 |
| Mannitol | 35.8 (41) | 24.4 | 32.6 (41) | 19.7 | 34.3 (41) | 23.7 | 0.46 |
| Sucralose | 3.2 (41) | 2.18 | 3.21 (41) | 2.59 | 3.08 (41) | 2.28 | 0.38 |
| Sucralose/Rhamnose | 0.47 (41) | 0.15 | 0.42 (41) | 0.19 | 0.46 (41) | 0.16 | 0.61 |
| Sucralose/Mannitol | 0.09 (41) | 0.04 | 0.09 (41) | 0.04 | 0.09 (41) | 0.05 | 0.37 |
| **Timepoint 2 (2-8 hrs)** | | | | | | | |
| Rhamnose | 10.6 (38) | 9.44 | 12.8(38) | 15.6 | 12.1 (38) | 10.6 | 0.22 |
| Mannitol | 51.8 (38) | 32.7 | 54.9 (38) | 61.8 | 50.3 (38) | 53.2 | 0.27 |
| Sucralose | 8.54 (38) | 9.26 | 9.62 (38) | 13.04 | 7.93 (38) | 9.61 | 0.54 |
| **Timepoint 3 (8-24 hrs)** | | | | | | | |
| Rhamnose | 7.73 (39) | 3.7 | 7.73 (39) | 4.73 | 7.63 (39) | 5.46 | 0.66 |
| Mannitol | 38.3 (39) | 55.8 | 43.2 (39) | 60.5 | 36.9 (39) | 63.7 | 0.58 |
| Sucralose | 4.92 (39) | 3.71 | 4.58 (39) | 2.58 | 4.61 (39) | 3.75 | 0.58 |
| Sucralose/Rhamnose | 0.66 (39) | 0.47 | 0.67 (39) | 0.31 | 0.72 (39) | 0.37 | 0.94 |
| Sucralose/Mannitol | 0.12 (39) | 0.11 | 0.13 (39) | 0.14 | 0.14 (39) | 0.12 | 0.3 |
| **Timepoint 4 (0-24 hrs)** | | | | | | | |
| Rhamnose | 30.6 (37) | 16.6 | 27.1 (37) | 24 | 27.3 (37) | 16.5 | 0.31 |
| Mannitol | 155.3 (37) | 82.5 | 141.1 (37) | 126.9 | 127.8 (37) | 90.7 | 0.08 |
| Sucralose | 19.6 (37) | 8.07 | 17.5 (37) | 17.6 | 16.8 (37) | 14 | 0.31 |

*Urinary sugar excretion (mg) and disaccharide to monosaccharide ratio at each dietary condition across different timepoints of urine collection.* Data are presented as median with sample size (n) in parenthesis and interquartile range (IQR). Statistical analysis was performed using the Friedman nonparametric test.

Title: *Persea americana* for Total Health (PATH-2): Effects of Avocado Consumption on Gastrointestinal Health in a Randomized, Crossover, Complete Feeding Trial

First author: María G. Sanabria-Véaz, R.D.

### Figures

*Supplemental Figure 1. Study design and sample collection timeline*

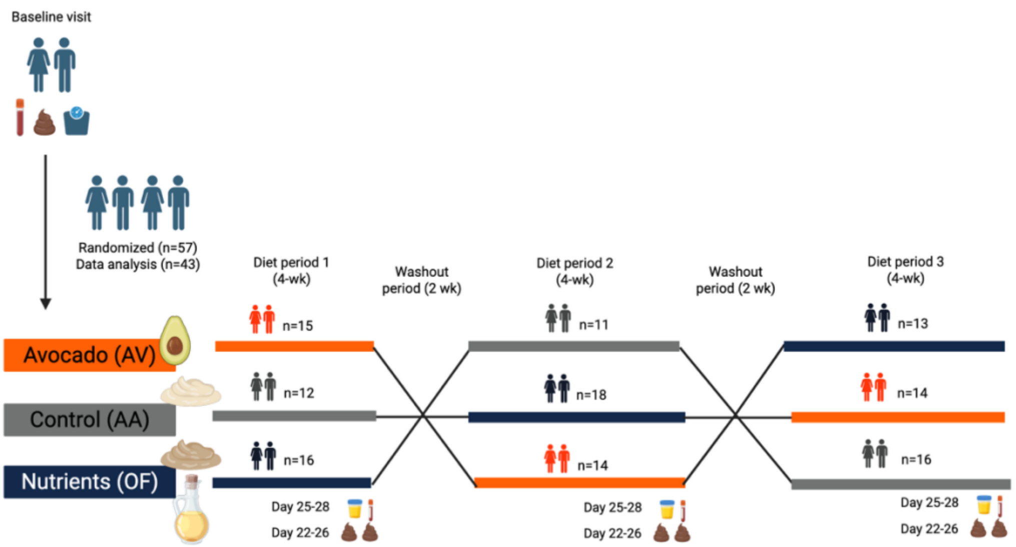


Participants were assigned to one of six treatment sequences (ABC, ACB, BAC, BCA, CAB, CBA) in a three-period crossover design, allowing each participant to serve as their own control. The three conditions included the Average American diet (AA), an oil-and-fiber–matched nutrients diet (OF), and an avocado diet (AV). Sample size (n) corresponds to participants among the 43 who completed the diet. Participants completed each 4-week condition, separated by 2-week washout periods, and provided fecal, blood, and urine samples at the end of each phase. *(Figure made with BioRender)*

Title: *Persea americana* for Total Health (PATH-2): Effects of Avocado Consumption on Gastrointestinal Health in a Randomized, Crossover, Complete Feeding Trial

First author: María G. Sanabria-Véaz, R.D.

*Supplemental Figure 2. Fecal calprotectin (A) and secretory IgA (B) levels across dietary conditions*

A.                                                    B.

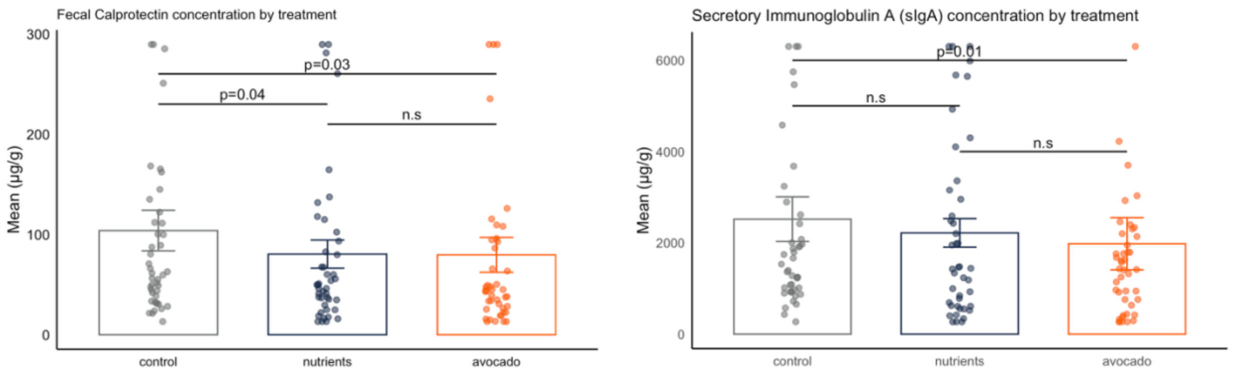


*Fecal calprotectin (A) and secretory IgA (B) levels across dietary conditions* Fecal calprotectin (A) and secretory IgA (B) were measured using ELISAs. Variables were log and/or square root transformed prior to repeated measures ANOVA to meet normality and homogeneity of variance assumptions. Fecal calprotectin and sIgA data points were winsorized at the 95th percentile for data visualization purposes only. Control = AA; Nutrients = OF; Avocado = AV
